# MOLECULAR ANALYSIS OF MITOCHONDRIAL COMPLEX I IN THE RESPONSIVENESS TO LEVODOPA IN PARKINSON’S DISEASE

**DOI:** 10.1101/2025.11.10.25339914

**Authors:** Ana Gabrielle Bispo, Felipe Gouvêa de Souza, Matheus Epifane-de-Assunção, Camille Sena-dos-Santos, Gustavo Barra Matos, Dafne Dalledone Moura, Gracivane Lopes Eufraseo, Caio Santos Silva, Gilderlanio Santana de Araújo, Ândrea Ribeiro-dos-Santos, Bruno Lopes Santos-Lobato, Giovanna C. Cavalcante

## Abstract

**Introduction:** Significant advances have been made in elucidating the pathophysiological mechanisms of Parkinson’s disease (PD). Levodopa remains the main therapeutic option — although it presents heterogeneous clinical benefits among patients. Mutations related to levodopa metabolic pathways have been investigated, but not for mtDNA. Since levodopa metabolism is highly dependent on ionic gradients, endocytosis, and vesicular transport — all ATP-dependent processes — the normal function of OXPHOS is essential not only for adequate levodopa metabolism but also for its therapeutic efficacy. This study aimed to analyze levodopa responsiveness profiles considering the mitochondrial genomic component in Brazilian admixed populations.

**Methods:** A total of 49 patients with PD underwent a levodopa challenge test (LCT), followed by whole mitochondrial genome sequencing, pathogenicity prediction of identified variants, and *in silico* structural analyses.

**Results:** Variants most frequently affected ND4, ND5, and ND6 subunits in both groups (responsive and non-responsive). Among them, the responsive group presented variants in *MT-ND4* (m.12018C>G - T420S) and ND5 (m.13130C>A - P265H) as those with the most significant structural impact, suggesting a loss of the native conformation and alterations in protein efficiency. Additionally, five unique variants were detected only among non-responsive patients, two of which were absent from the dbSNP and ClinVar databases, which indicates the possibility that they are novel variants and potentially population-specific.

**Conclusion:** We provide molecular evidence suggesting that variants in mitochondrial ND4, ND5, and ND6 subunits, in addition to mitochondrial ancestry, may contribute to distinct levodopa responsiveness in PD patients.

## 1. INTRODUCTION

Parkinson’s disease (PD) is a progressive neurodegenerative condition that primarily affects dopaminergic neurons of the nigrostriatal pathway [1]. Consequently, motor decline represents one of the most prominent phenotypes of the disease, characterized by resting tremor, postural instability, bradykinesia, and muscle rigidity [2]. Among the available treatment strategies, Levodopa remains the most widely used and effective medication against motor symptoms. Levodopa is a metabolic precursor that, upon crossing the blood– brain barrier, is converted into dopamine in the remaining neurons, thereby restoring motor function [3]. Responsiveness to levodopa is subject to wide interindividual variability, accompanied by a progressive decline in pharmacological benefit as the disease progresses, necessitating ongoing adjustments in therapeutic management [4,5].

As underlying factors of this heterogeneous responsiveness, genetic variability in pharmacokinetic and pharmacodynamic pathways has been investigated [6]. Moreover, mitonuclear genomic ancestry has been shown to play a significant role in drug response and susceptibility to adverse reactions [6,7]. Mutations associated with mitochondrial dysfunction also appear to be essential contributors previously explored in the context of pathogenesis, though they remain not investigated with respect to the modulation of PD treatment. Because levodopa metabolism further challenges mitochondrial function, individual mitochondrial genetic backgrounds may modulate therapeutic response and susceptibility to treatment-related complications, highlighting the importance of integrating mitochondrial genetics into the understanding of pharmacological variability in PD.

Among their several functions, mitochondria generate the majority of cellular ATP through the five protein complexes of oxidative phosphorylation (OXPHOS) [8]. Some studies worldwide have investigated levodopa and Complex I (CI), suggesting that CI inhibition due to defects in its core subunits may be related to PD progression and responsiveness to Levodopa, as such impairments can disrupt ATP production, increase oxidative stress, and trigger neuronal vulnerability [9, 10, 11].

In addition, a significant limitation identified in research on levodopa responsiveness concerns the lack of interethnic genomic representativeness [7]. In this context, Brazil has a population with diverse genomic ancestry contributions, but there is still limited reported data on pharmacogenetics, making the Brazilian population a relevant focus for investigating interindividual variation in levodopa responsiveness.

Therefore, the present study adopted the Levodopa Challenge Test (LCT) alongside the investigation of genomic ancestry and genetic alterations in the mitogenome that may influence treatment response and lead to motor phenotypes. This approach aims to expand knowledge of the mitochondrial genetic influence on PD therapy and to improve understanding of individual variations in drug efficacy, with the ultimate goal of guiding the future development of more efficient and individualized therapeutic strategies, thereby supporting patient autonomy, functional mobility, and, consequently, quality of life.

## 2. METHODS

### 2.1. Sampling and ethical aspects

For this study, we recruited people with PD from the Movement Disorders Unit of the Hospital Ophir Loyola, Brazil, who are treated with Levodopa. The following inclusion criteria were applied: (i) individuals diagnosed with idiopathic Parkinson’s disease according to the London Brain Bank diagnostic criteria; and (ii) patients who had been using L-DOPA for at least three months. The exclusion criteria were as follows: (i) inability to adequately complete the questionnaires; (ii) presence of disabling neurological disorders, such as other movement disorders, neurodegenerative diseases, epilepsies, neuroimmunological diseases, infections, or neoplasms of central nervous system; (iii) presence of severe and disabling non-neurological diseases (e.g., malignant neoplasms, severe cardiac, hepatic or renal failure); (iv) prior neurosurgical procedures for the treatment of Parkinson’s disease (deep brain stimulation or ablative surgery); (v) history of significant gastroduodenal disorders (e.g., gastric or duodenal ulcers, gastric cancer) or gastric surgeries; (iv) use of antibiotics within last four weeks; and (vii) use of antacids, H_2_ receptor antagonists, proton pump inhibitors, domperidone, prokinetic agents, or any other drugs affecting gastrointestinal motility within the last two weeks. For individuals who had been using L-DOPA for less than three months, antibiotics for more than four weeks, and/or other medications for more than two weeks, recruitment was postponed, and specific instructions were provided.

Afterwards, participants were categorized into two subgroups: people with PD who had a good response to levodopa (responders) and those who had a poor response (non-responders) (Figure 1). The project was submitted and approved by the Research Ethics Committee of the Ophir Loyola Hospital (CAAE 62518222.7).

**Figure 1.**
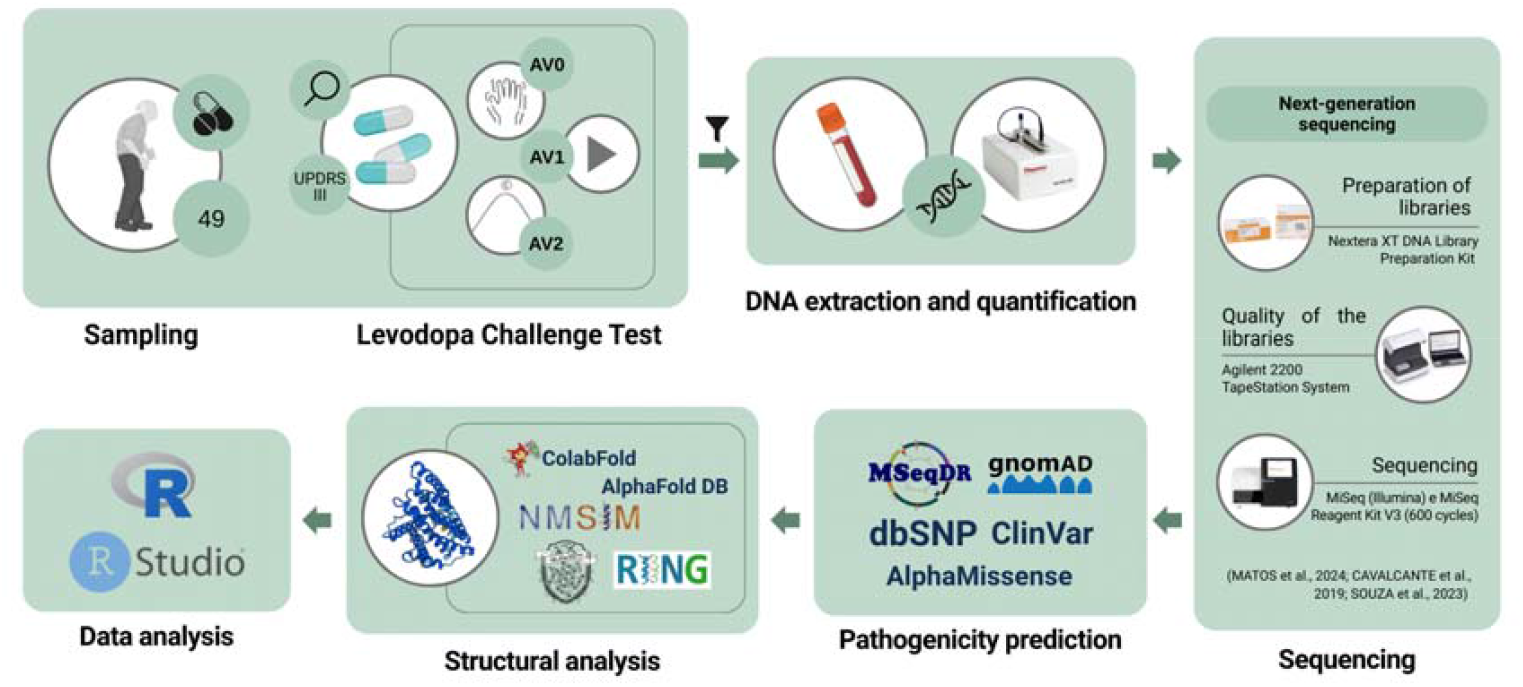
Workflow representing the methodological approach adopted.

The study began with a sample of 49 PD patients diagnosed by a clinical specialist who underwent the Levodopa challenge test, with motor assessment based on the UPDRS scale. Part of the initial sample was forwarded to the next steps, which included DNA extraction and quantification, followed by next-generation sequencing of the mitochondrial genome. The CI variants were assessed for pathogenicity using databases, followed by a structural evaluation of the mutant proteins.

### 2.2. Levodopa responsiveness

To assess patients’ response to the medication, the LCT [12, 13] was performed, which is conducted based on the Movement Disorders Society - Unified Parkinson’s Disease Rating Scale (MDS-UPDRS) Part III [14]. The MDS-UPDRS Part III score allows the analysis of subscores, which correspond to the variation in patients’ motor symptoms calculated from the difference between the assessment scores. These subscores are: tremor (including items assessing postural and kinetic tremor of the hands, amplitude and persistence of resting tremor), rigidity (referring to the upper and lower limbs and neck), bradykinesia (facial expression, finger tapping, hand opening and closing movement, hand pronation-supination, toe tapping, leg agility and global spontaneity of movement) and axial symptoms (based on speech, gait, gait blockage - freezing, postural stability and posture) [14, 15].

The same motor aspects were assessed at three moments: Period A (“off” state), Period B (onset of effect), and Period C (peak dose), which represent the “on” state. Period A or AV0 is assessed with the patient having not taken Levodopa or any other antiparkinsonian drug for at least 12 hours. Then, levodopa is administered, usually at 150% of the usual dose, and after 20 minutes or upon the patient’s subjective report regarding the onset of effect, new assessments are performed (Periods B or AV1 and C or AV2). It is important to note that all assessments are filmed and subsequently analyzed by a specialized neurologist (author B.L.S.L). Finally, the formula for the percentage of response to levodopa (%RL) is applied, %RL = (UPDRS AV0 - UPDRS AV2/UPDRS AV0), and the cut-off value that indicates significant response capacity to Levodopa is greater than or equal to 33% [16].

### 2.3. DNA extraction and quantification

The genetic analysis of the present study was based on mitogenome sequencing previously conducted by the research group [17]. In this stage of the project, 15 individuals were included, 12 from the responsive group and three from the non-responsive group. DNA was extracted from the peripheral blood samples of each patient using the phenol-chloroform method, as described by Sambrook et al. [18]. Quantification of total DNA was measured with the NanoDrop 1000 spectrophotometer (Thermo Fisher Scientific, Wilmington, DE, USA).

### 2.4. Sequencing

For amplification of the mitochondrial genome, the primers previously designed [19] were used in the established PCR protocol [17]. To construct the libraries from the amplified material, the Nextera XT DNA Library Preparation Kit (Illumina Inc., Chicago, IL, USA) was used, and the MiSeq Reagent Kit V3 (600 cycles) (Illumina) was used for sequencing on the MiSeq system (Illumina), according to the manufacturer’s instructions. To prepare the libraries, the High Sensitivity D1000 ScreenTape was also used on the Agilent 2200 TapeStation System (Agilent Technologies, Santa Clara, CA, USA) to assess the quality of the libraries obtained. After sequencing was completed, a specific pipeline was adopted for bioinformatic preparation and analysis, aligned with the revised Cambridge reference sequence, as previously established [19, 20]. Then, the mitochondrial regions of interest (CI genes and D-loop region) were selected for further analysis.

### 2.5. Pathogenicity prediction

The genomic consequence, impact, and pathogenicity prediction were inferred from the Mitochondrial Disease Sequence Data Resource (MSeqDR, https://mseqdr.org/index.php) platform [21]. In addition, the following databases were used: genome aggregation (gnomAD, https://gnomad.broadinstitute.org/) [22], single nucleotide polymorphism (dbSNP, https://www.ncbi.nlm.nih.gov/snp/) [23], and ClinVar (https://www.ncbi.nlm.nih.gov/clinvar/) [24] for clinical consequence analysis. Additionally, the analysis was incorporated into AlphaMissense (https://alphamissense.hegelab.org/) [25], a prediction tool, to assess the possible functional impact of mutations in proteins.

### 2.6. Structural analysis

To analyze the structural impact of mutations, mutant proteins were modeled using ColabFold [26]. Wild-type models were obtained directly from AlphaFoldDB, using its code in the revised UniProt [27, 28].

The wild-type and mutant models obtained were subjected to a geometric approximation simulation of molecular dynamics using the NMSIM tool [29]. Using this tool, 500 conformational variants were generated for each protein studied, representing the structural variations that the protein could assume in three-dimensional (3D) space based on its amino acid composition and physical-chemical characteristics. The 3D visualization of the wild-type and mutant trajectories was performed using the VMD (Visual Molecular Dynamics) tool [30].

From these trajectory data, a comparison of the Root Mean Square Deviation (RMSD) of the wild-type and mutant proteins was performed. In addition, the Root Mean Square Fluctuation (RMSF) values were also compared, which allows us to individually investigate whether the dynamic performance of each residue of the wild-type protein was altered by undergoing the mutation. The generated files, containing the conformational variants, also known as trajectory files, were processed by the RING (Residue Interaction Network Generator) tool to construct the residue interaction networks of the proteins [31]. Finally, the comparative analysis of the residue interaction networks of the conformational variants of the wild-type and mutant proteins was performed using the SlytheRINs tool (https://github.com/jpmslima/SlytheRINs.git), in order to verify changes in the binding patterns to provide insights into possible conformational transitions resulting from mutations in structural flexibility and/or rigidity.

### 2.7. Data analysis

All statistical analyses were performed in R [32] and visualized in RStudio [33]. For graphical representations and statistical analyses performed in R, the following packages were used: ggplot2 [34], rstatix [35], UpSetR [36], ggpubr [37], and ggcorrplot [38], with the Wilcoxon, Student’s t, Spearman, and chi-squared tests being used when appropriate. For all statistical tests, p < 0.05 was considered statistically significant.

## 3. RESULTS

### 3.1. Clinical aspects of the cohort

In this study, the cohort consisted of 49 individuals with PD. We observed similar mean ages between the groups and no statistically significant differences in the sex distribution, percentage of dyskinesia, family history, initial symptoms, or other clinical characteristics investigated (Table 1).

**Table 1.**
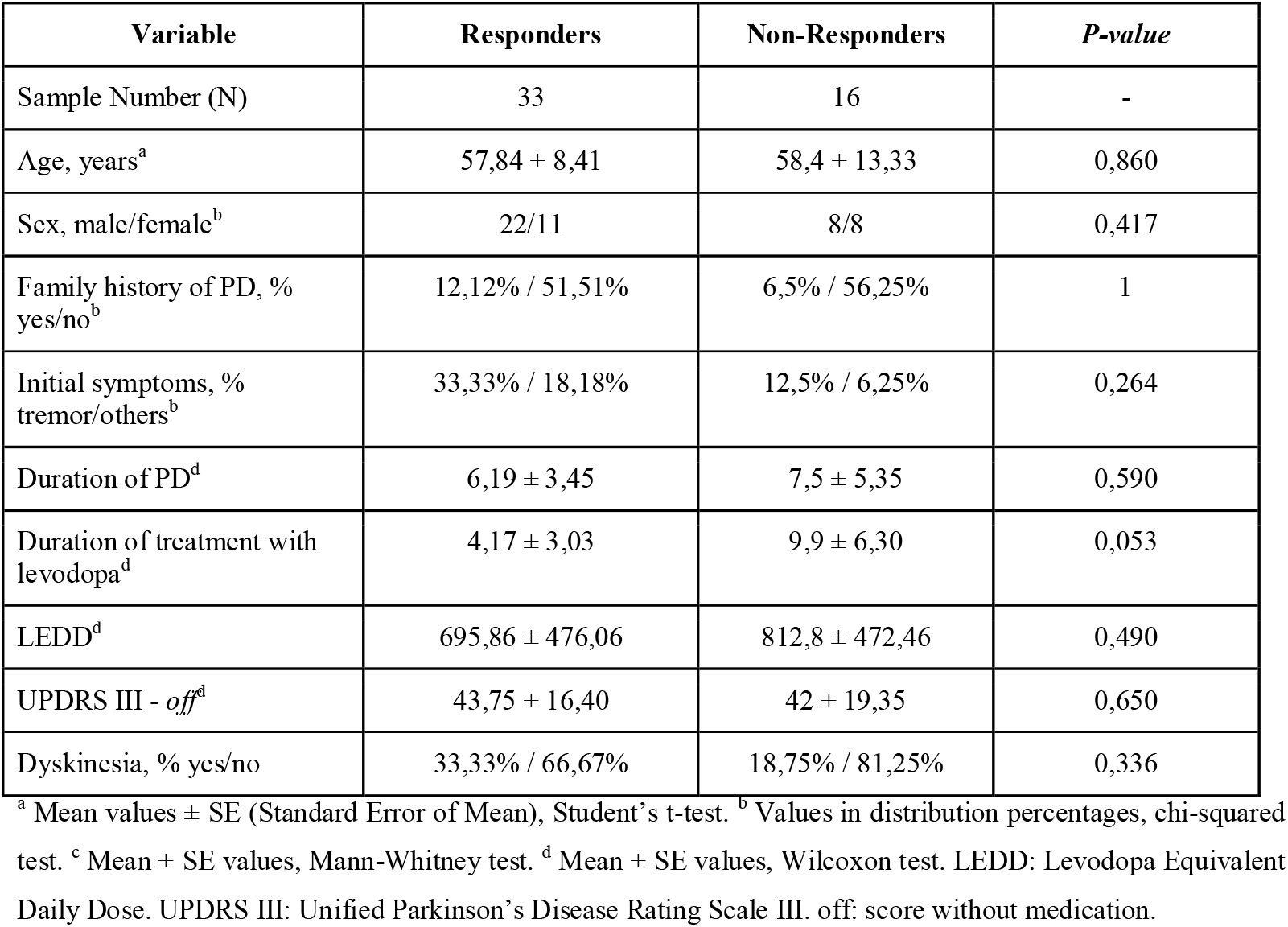
Demographic characteristics of the groups included in the study.

| Variable | Responders | Non-Responders | *P-value* |
| --- | --- | --- | --- |
| Sample Number (N) | 33 | 16 | - |
| Age, years<sup>a</sup> | 57,84 ± 8,41 | 58,4 ± 13,33 | 0,860 |
| Sex, male/female<sup>b</sup> | 22/11 | 8/8 | 0,417 |
| Family history of PD, % yes/no<sup>b</sup> | 12,12% / 51,51% | 6,5% / 56,25% | 1 |
| Initial symptoms, % tremor/others<sup>b</sup> | 33,33% / 18,18% | 12,5% / 6,25% | 0,264 |
| Duration of PD<sup>d</sup> | 6,19 ± 3,45 | 7,5 ± 5,35 | 0,590 |
| Duration of treatment with levodopa<sup>d</sup> | 4,17 ± 3,03 | 9,9 ± 6,30 | 0,053 |
| LEDD<sup>d</sup> | 695,86 ± 476,06 | 812,8 ± 472,46 | 0,490 |
| UPDRS III - *off*<sup>d</sup> | 43,75 ± 16,40 | 42 ± 19,35 | 0,650 |
| Dyskinesia, % yes/no | 33,33% / 66,67% | 18,75% / 81,25% | 0,336 |

<sup>a</sup> Mean values ± SE (Standard Error of Mean), Student's t-test. <sup>b</sup> Values in distribution percentages, chi-squared test. <sup>c</sup> Mean ± SE values, Mann-Whitney test. <sup>d</sup> Mean ± SE values, Wilcoxon test. LEDD: Levodopa Equivalent Daily Dose. UPDRS III: Unified Parkinson's Disease Rating Scale III. *off*: score without medication.

### 3.2. Responsiveness to levodopa

Figure S1 shows the comparison of the variations of these subscores obtained by the MDS-UPDRS III between responsive and non-responsive individuals, stratified by sex. Overall, it is observed that both sexes in the Responsive group showed improvement in all subscores analyzed and that, for symptoms such as tremor, bradykinesia, and rigidity, the differences between sexes were not statistically significant for either group. However, in Figure S1D, a statistically significant difference between the genders in the variation of axial symptoms in the responsive group (p = 0.03) is notable, suggesting that women who are responsive to LD may exhibit a greater variation in axial symptoms between the motor assessments before and after LD administration.

Spearman’s correlation analysis, presented in Figure S2, revealed significant associations between the responsiveness and the motor subscores. In the total sample, the LCT result showed a strong positive correlation with the variation in bradykinesia (r = 0.9; p-adjust= 1.963973e-14), in addition to moderate correlations with the symptoms of tremor (r = 0.6; p-adjust= 5.577845e-05), rigidity (r = 0.4; p-adjust= 4.076954e-02), and axial symptoms (r = 0.6; p-adjust= 5.810891e-05). In the responsive group, bradykinesia remained the symptom most associated with the response to LCT (r = 0.8; p-adjust= 1.349498e-08), followed by axial symptoms (r = 0.5; p-adjust= 0.002245238). In the non-responsive group, the correlations were lower, with only bradykinesia standing out (r = 0.6; p-adjust= 0.047477776). In all groups, sex did not show significant correlations with the clinical variables.

### 3.3. Genetic ancestry and levodopa responsiveness

Nuclear ancestry showed a statistical difference in both LCT responsiveness groups, especially considering European ancestry in relation to other ancestries, indicating that the largest proportion is of European nuclear ancestry (Figure 2A). Despite this, we observed that Native American mitochondrial ancestry is more frequent in the cohort, specifically the macrohaplogroup C in the responsive group (Figure 2B).

**Figure 2.**
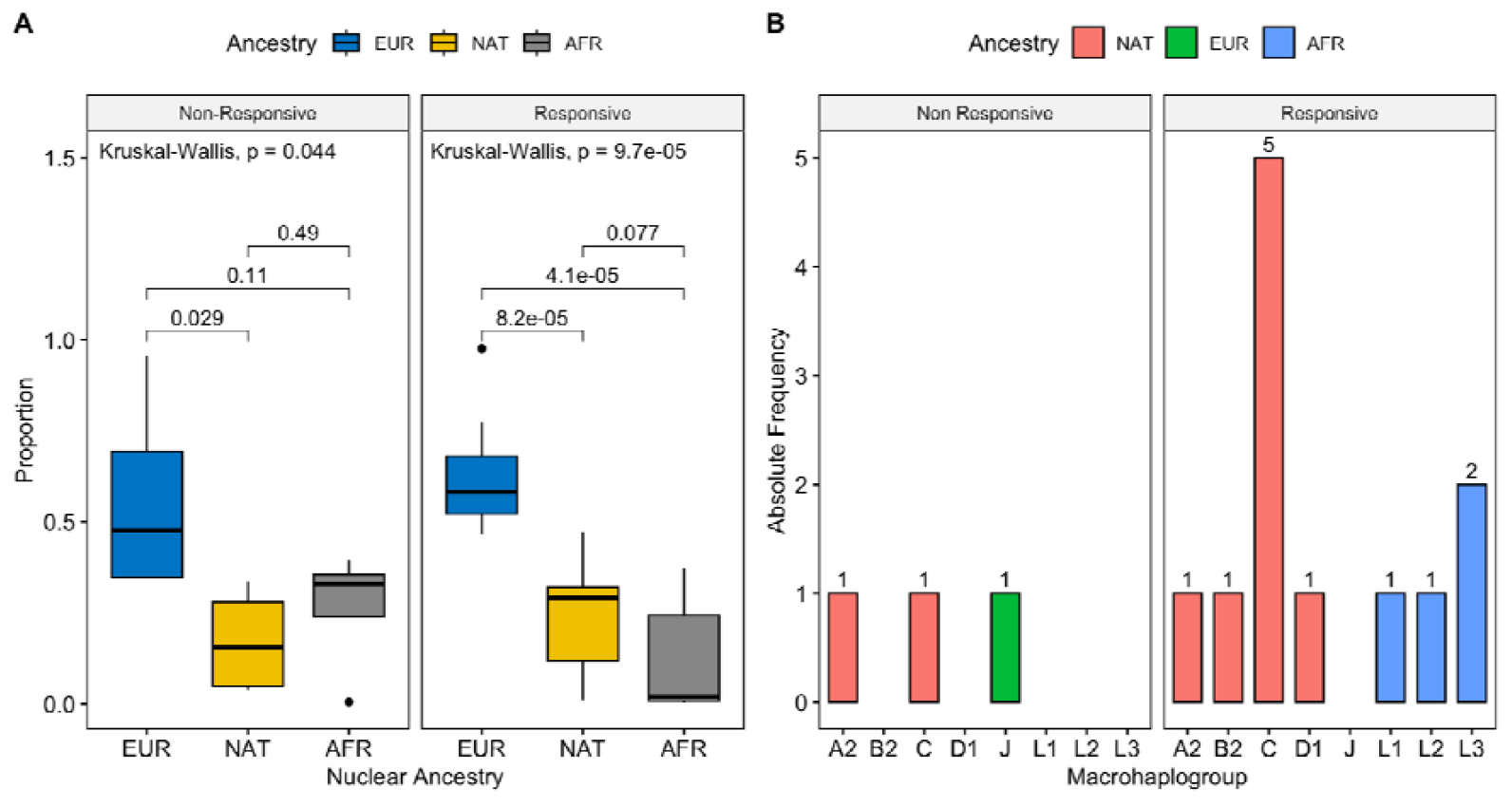
Genomic ancestry of the LCT-responsive and non-responsive patient groups. A: Nuclear ancestry of the LCT-responsive and non-responsive patient groups. B: Mitochondrial ancestry of the LCT-responsive and non-responsive patient groups. EUR: European ancestry. NAT: Native American or indigenous ancestry. AFR: African ancestry.

### 3.4. Mutation profiling

After normalizing the number of variants by gene length, we found a greater presence of mutations in the *MT-ND5* gene in relation to the others in both groups, with the responsive group having approximately three times more unique mutations than the non-responsive group (Figure S3).

When analyzing the absolute frequency of mutation types in terms of genomic consequence based on the gnomAD database, it is possible to observe that most of the mutations found are synonymous, with a much higher number in *MT-ND5* in both responsiveness groups (Figures 3A and 3C). From the AlphaMissense pathogenicity predictor, it is noted that, in both responsiveness groups, there are mutations inferred as probably pathogenic, ambiguous and probably benign, and the only genes that presented mutations with one of the three classifications were *MT-ND4* and *MT-ND5* (Figures 3B and 3D). These genes encode essential subunits for the formation of the NADH dehydrogenase respiratory chain in the mitochondrial inner membrane, thus likely playing a critical role in OXPHOS [42].

**Figure 3.**
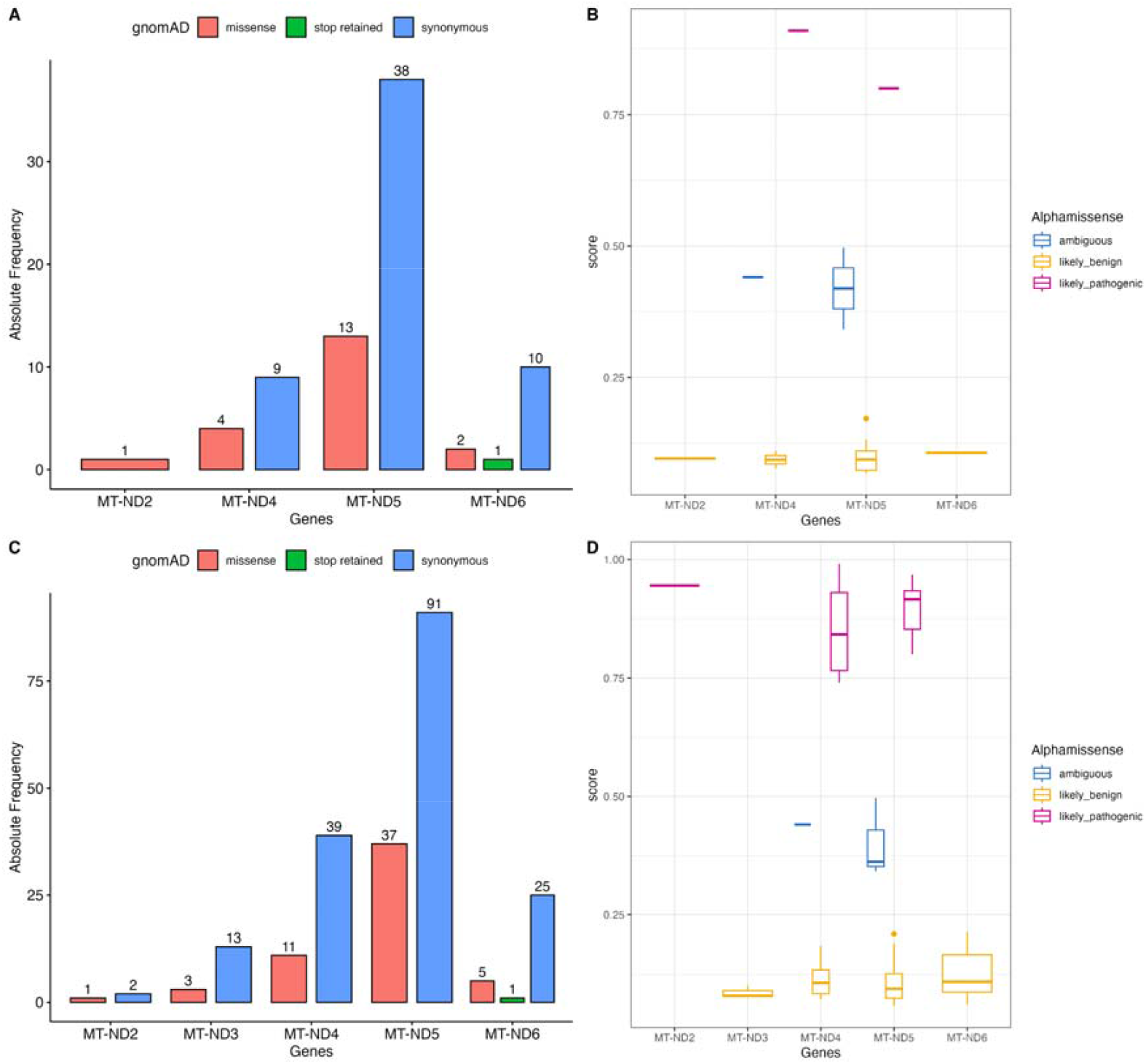
Distribution of heteroplasmic variants per gene according to LCT responsiveness groups with their predicted consequences. A: Genomic consequence of mitochondrial variants encoding OXPHOS complex I subunits from LCT-unresponsive patients. B: Pathogenicity score of missense variants from LCT-responsive patients inferred by AlphaMissense. C: Genomic consequence of mitochondrial variants encoding OXPHOS complex I subunits from LCT-responsive patients. D: Pathogenicity score of missense variants from LCT-responsive patients inferred by AlphaMissense.

In the analysis of the levels of heteroplasmy, no statistically significant difference was observed between the LCT-responsive and non-responsive groups. However, when considering the comparisons between genes within each group, relevant differences were observed, as shown in Figure S4. In the non-responsive group, the comparisons between the *MT-ND4* and *MT-ND5* genes, as well as between *MT-ND5* and *MT-ND6*, revealed statistical significance (Figure S4A). In the responsive group, the *MT-ND5* gene presented significantly higher levels of heteroplasmy in relation to most of the other genes analyzed (Figure S4B). Consistently between both groups, *MT-ND5* was the gene that presented the highest levels of heteroplasmy within CI.

In the Supplementary Material, Table S1 shows the most frequent CI mutations in the responsive group, considering those present in at least 60% of the cohort of patients with responsiveness in the LCT, but not necessarily exclusive to this group. It should be noted that only the *MT-ND4* and *MT-ND5* genes appear in this list, which may be related to the greater number of mutations in these genes, and that three and two mutations, respectively, are of the missense type, while the others are synonymous. Furthermore, the missense mutations 12018C>G - T420S (*MT-ND4*), 13130C>A - P265H and 13130C>T - P265L (*MT-ND5*) were not reported in any genomic and clinical database, having their predictions in AlphaMissense as probably benign (12018C>G - T420S) and ambiguous (13130C>A - P265H and 13130C>T - P265L).

Figure S5 shows the number of mutations unique to each LCT-responsiveness group and the number of mutations shared by both groups. The characterization of these unique and shared mutations can be seen in Tables S2-S4. A total of 73 shared mutations were identified, of which 18 are missense variants (Table S4), most of which do not present current records in the dbSNP and ClinVar databases. Regarding the variants exclusive to each group, 155 variants were identified in the responsive group, of which four synonymous variants were present in at least 60% of the cohort, located in the *MT-ND4* and *MT-ND5* genes (Table S3). In the non-responsive group, five exclusive variants were identified (Table S2), located in the *MT-ND2, MT-ND4, MT-ND5*, and *MT-ND6* genes. Among them, the missense mutations 5322A>C (*MT-ND2*) and 14170A>T (*MT-ND6*) stand out, which are not currently recorded in the dbSNP and ClinVar databases, suggesting these might be novel variants with unfavorable potential for responsiveness to L-DOPA.

### 3.5. Structural analysis of mutant proteins

Next, structural analysis was performed from the seven CI missense mutations found among the most frequent (present in at least 60% of the cohort) in patients with LCT responsiveness (*MT-ND4* (UniProt ID: P03905): 11505T>C - I249T, 11963G>A - V402I, 12018C>G - T420S; *MT-ND5* (UniProt ID: P03915): 13130C>A - P265H, 13130C>T - P265L) or exclusive to patients with low LCT responsiveness (*MT-ND2* (UniProt ID: P03891): 5322A>C - I285L; *MT-ND6* (UniProt ID: P03923): 14170A>T - I168M). These variants are described in Tables S2 and S3 of the Supplementary Material. Videos of the dynamic trajectories of wild-type and mutant proteins are available in the repository: https://github.com/matheus4ssuncao/Bispo-et-al-2025.

Among the investigated mutations, I249T, P265H, and I168M showed relevant discrepancies in the global structure analysis based on RMSD when compared to their WT forms, with P265H standing out due to greater global destabilization. Regarding RMSF values, I249T exhibited increased flexibility, mainly in its first 200 residues, while I168M resulted in a decrease in flexibility. V402I, P265L, and I285L did not show substantial variations in the RMSD and RMSF comparisons. Similarly, T420S and P265H displayed greater variability in residue-residue interactions, suggesting increased disruption of internal bonds and reduced local stability. As for the interaction analysis of the other variants, no significant pattern was observed (Figures S6–12).

## 4. DISCUSSION

We observed improvement in all motor symptoms in the responsive group, as also demonstrated by the strong positive correlation between LCT and bradykinesia, and a moderate correlation for the other symptoms. Bradykinesia was associated with LCT in both groups, suggesting that this symptom may be the most sensitive to L-DOPA effects, even in individuals showing suboptimal treatment response (10.1038/s41531-022-00321-y).

Among the motor features, axial symptoms are notable for their gender-related differences, with female individuals exhibiting a statistically significant clinical benefit compared to their male counterparts. This finding is consistent with studies indicating that women have greater levodopa bioavailability than men. The pharmacokinetics and pharmacodynamics of levodopa in female patients may be mainly influenced by factors such as lower body weight, slower gastric emptying time, higher plasma levodopa concentrations, reduced clearance, and lower COMT enzymatic activity [39–41], allowing them to experience more of the drug’s pharmacological benefits.

In terms of ancestry, European nuclear ancestry predominated across the cohort, whereas Native American ancestry showed higher overall prevalence, with macrohaplogroup C being more frequent among responsive individuals. These observations reflect the processes of occupation, migration, and colonization in the northern territory of Brazil, as described in other works by the research group on this population [19, 20].

The responsive group presented three times more SNVs than the non-responsive group, along with a higher heteroplasmy level in the MT-ND5 gene compared to the other genes in the group, suggesting a possible central role of the observed mitochondrial alterations in treatment responsiveness. To further elucidate the molecular mechanisms underlying the role of these SNVs in modulating responsiveness, a structural investigation of the missense variants was performed.

That said, the T420S variant was one of those investigated. It affects the ND4 subunit of CI, which forms the distal portion of the hydrophobic arm of the complex [43]. This subunit is believed to act as an antiporter-like protein and therefore contributes to the formation of the electrochemical gradient required for ATP synthesis [44, 45]. Although it does not constitute a transmembrane segment — structures traditionally more associated with proton translocation — some studies provide evidence that residues located outside the transmembrane helices, from residue 400 onwards, can also influence the proton-pumping mechanism [46]. The study by TORRES-BACETE et al. [47] shows that the C-terminal region of the bacterial homologue of ND4 (NuoM) is essential for enzyme activity, demonstrating a partial loss of NADH:quinone oxidoreductase function following truncations in different C-terminal segments.

Similar impacts can be observed for mutations in ND5 subunit, resulting in elevated levels of reactive oxygen species (ROS), altered mitochondrial membrane potential, ATP depletion and reduced cell viability [51, 52, 53]. P265H is an unreported variant. Although no studies have investigated the structural and functional impact of this specific mutation, a variant at position 236 of ND5 has been associated with mitochondrial and energy metabolism dysfunction. [54].

Although studies in the literature that investigated nearby mutations generally report some degree of CI deficiency and impairment of the cellular physiological phenotype [52, 53, 54], our data indicate that this conformational change in the ND5 subunit may favor mitochondrial bioenergetics and improve the response to levodopa.

## 5. CONCLUSIONS

We provide molecular evidence suggesting that variants in *MT-ND4, MT-ND5*, and *MT-ND6* contribute to distinct levodopa responsiveness in PD patients. Structural analyses suggest that some variants in responders may favorably influence OXPHOS function without causing CI dysfunction. The non-responsive group harbored unique variants, including two novel ones. Differences in genomic ancestry were also observed, with higher Native American mitochondrial ancestry (notably macrohaplogroup C) among responders. These findings highlight the role of mitochondrial genetics and ancestry in modulating therapeutic response in PD and point to the potential for population-specific biomarkers.

## Data Availability

All data produced in the present work are contained in the manuscript.

## Data Availability

Supplementary Material is available at the journal’s website. The analyzed data are available at the European Nucleotide Archive (ENA) under the accession number PRJEB74357. Videos of the dynamic trajectories are available in the repository: https://github.com/matheus4ssuncao/Bispo-et-al-2025.

## CRediT authorship contribution statement

**AGB:** Conceptualization, Methodology, Investigation, Writing - Original Draft, Writing - Review & Editing, Visualization. **FGS:** Methodology, Formal analysis, Investigation, Writing - Original Draft, Writing - Review & Editing, Visualization. **ME-d-A:** Methodology, Formal analysis, Investigation, Writing - Original Draft, Writing - Review & Editing, Visualization. **CS-d-S:** Methodology. **GBM:** Methodology. **DDM:** Data curation. **GLE:** Data curation. **CSS:** Methodology. **GSA:** Methodology, Supervision. **ÂR-d-S:** Resources, Funding acquisition, Project administration. **BLSL:** Resources, Data curation, Funding acquisition, Project administration. **GCC:** Conceptualization, Investigation, Writing - Original Draft, Writing - Review & Editing, Visualization, Supervision, Project administration.

## Ethics approval

The project was approved by the Research Ethics Committee of the Ophir Loyola Hospital (CAAE 62518222.7), Brazil.

## Consent to participate

Informed consent was obtained from all participants in the study.

## Acknowledgements

The authors thank the participants of the study and the funding agencies.

## REFERENCES

[1] D.W. Dickson, Neuropathology of Parkinson disease, Parkinsonism & Related Disorders. 46 (2018) S30–S33. 10.1016/j.parkreldis.2017.07.033.

[2] D. Aarsland, L. Batzu, G.M. Halliday, G.J. Geurtsen, C. Ballard, K. Ray Chaudhuri, D. Weintraub, Parkinson disease-associated cognitive impairment, Nature Reviews Disease Primers. 7 (2021). 10.1038/s41572-021-00280-3.

[3] J. Jankovic, E.K. Tan, Parkinson’s Disease: Etiopathogenesis and Treatment, Journal of Neurology, Neurosurgery & Psychiatry. 91 (2020) 795–808. 10.1136/jnnp-2019-322338.

[4] C. Strafella, V. Caputo, M.R. Galota, S. Zampatti, G. Marella, S. Mauriello, R. Cascella, E. Giardina, Application of Precision Medicine in Neurodegenerative Diseases, Frontiers in Neurology. 9 (2018). 10.3389/fneur.2018.00701.

[5] M. Beckers, B.R. Bloem, M.M. Verbeek, Mechanisms of peripheral levodopa resistance in Parkinson’s disease, Npj Parkinson’s Disease. 8 (2022). 10.1038/s41531-022-00321-y.

[6] E.U.D. Dos Santos, I.I.F.G. da Silva, A.G.C. Asano, N.M.J. Asano, M. De Mascena Diniz Maia, P.R.E. de Souza, Pharmacogenetic profile and the development of the dyskinesia induced by levodopa-therapy in Parkinson’s disease patients: a population-based cohort study, Molecular Biology Reports. 47 (2020) 8997–9004. 10.1007/s11033-020-05956-9.

[7] M. Bachtiar, B.N.S. Ooi, J. Wang, Y. Jin, T.W. Tan, S.S. Chong, C.G.L. Lee, Towards precision medicine: interrogating the human genome to identify drug pathways associated with potentially functional, population-differentiated polymorphisms, The Pharmacogenomics Journal. 19 (2019) 516–527. 10.1038/s41397-019-0096-y.

[8] W. Kühlbrandt, Structure and function of mitochondrial membrane protein complexes, BMC Biology. 13 (2015). 10.1186/s12915-015-0201-x.

[9] D. Ben-Shachar, R. Zuk, H. Gazawi, P. Ljubuncic, Dopamine toxicity involves mitochondrial complex I inhibition: implications to dopamine-related neuropsychiatric disorders, Biochemical Pharmacology. 67 (2004) 1965–1974. 10.1016/j.bcp.2004.02.015.

[10] N. Subrahmanian, M.J. LaVoie, Is there a special relationship between complex I activity and nigral neuronal loss in Parkinson’s disease? A critical reappraisal, Brain Research. (2021) 147434. 10.1016/j.brainres.2021.147434.

[11] S. Przedborski, V. Jackson-Lewis, U. Muthane, H. Jiang, M. Ferreira, A.B. Naini, S. Fahn, Chronic levodopa administration alters cerebral mitochondrial respiratory chain activity, Annals of Neurology. 34 (1993) 715–723. 10.1002/ana.410340515.

[12] G.L. Defer, H. Widner, R.M. Marié, P. Rémy, M. Levivier, Core assessment program for surgical interventional therapies in Parkinson’s disease (CAPSIT-PD), Movement Disorders: Official Journal of the Movement Disorder Society. 14 (1999) 572–584. 10.1002/1531-8257(199907)14:4%3C572::aid-mds1005%3E3.0.co;2-c.

[13] J.W. Langston, H. Widner, C.G. Goetz, D. Brooks, S. Fahn, T. Freeman, R. Watts, Core assessment program for intracerebral transplantations (CAPIT), Movement Disorders. 7 (1992) 2–13. 10.1002/mds.870070103.

[14] C.G. Goetz, B.C. Tilley, S.R. Shaftman, G.T. Stebbins, S. Fahn, P. Martinez-Martin, W. Poewe, C. Sampaio, M.B. Stern, R. Dodel, B. Dubois, R. Holloway, J. Jankovic, J. Kulisevsky, A.E. Lang, A. Lees, S. Leurgans, P.A. LeWitt, D. Nyenhuis, C.W. Olanow, Movement Disorder Society-sponsored revision of the Unified Parkinson’s Disease Rating Scale (MDS-UPDRS): Scale presentation and clinimetric testing results, Movement Disorders. 23 (2008) 2129–2170. 10.1002/mds.22340.

[15] X. Li, Y. Xing, A. Martin-Bastida, P. Piccini, D.P. Auer, Patterns of grey matter loss associated with motor subscores in early Parkinson’s disease, NeuroImage Clinical. 17 (2017) 498–504. 10.1016/j.nicl.2017.11.009.

[16] G. Saranza, A.E. Lang, Levodopa challenge test: indications, protocol, and guide, Journal of Neurology. 268 (2020). 10.1007/s00415-020-09810-7.

[17] G.B. Matos, C.S. Santos, T.P. Sousa, G. C Cavalcante, C.S. Silva, R.L.S. Cruz, D.D. Moura, A. Ribeiro-dos-Santos, B.L.S. Lobato, G.S. Araújo, The mitogenome mutation repertoire affects progression of Parkinson’s Disease, PREPRINT, Research Square. (2024). 10.21203/rs.3.rs-5411701/v1.

[18] J. Sambrook, E.F. Fritsch, T. Maniatis, Molecular Cloning: A Laboratory Manual, 2nd ed., Cold Spring Harbor Laboratory Press., 1989.

[19] G.C. Cavalcante, A.N.R. Marinho, A.K. Anaissi, T. Vinasco-Sandoval, A. Ribeiro-dos-Santos, A.F. Vidal, G.S. de Araújo, S. Demachki, Â. Ribeiro-dos-Santos, Whole mitochondrial genome sequencing highlights mitochondrial impact in gastric cancer, Scientific Reports. 9 (2019). 10.1038/s41598-019-51951-x.

[20] F.G. de Souza, M.B. da Silva, G.S. de Araújo, C.S. Silva, A.H.G. Pinheiro, M.Á. Cáceres-Durán, M.N. Santana-da-Silva, P. Pinto, A.R. Gobbo, P.F. da Costa, C.G. Salgado, Â. Ribeiro-dos-Santos, G.C. Cavalcante, Whole mitogenome sequencing uncovers a relation between mitochondrial heteroplasmy and leprosy severity, Human Genomics. 17 (2023). 10.1186/s40246-023-00555-8.

[21] M.J. Falk, L. Shen, M. Gonzalez, J. Leipzig, M.T. Lott, A.P.M. Stassen, M.A. Diroma, D. Navarro-Gomez, P. Yeske, R. Bai, R.G. Boles, V. Brilhante, D. Ralph, J.T. DaRe, R. Shelton, S.F. Terry, Z. Zhang, W.C. Copeland, M. van Oven, H. Prokisch, Mitochondrial Disease Sequence Data Resource (MSeqDR): A global grass-roots consortium to facilitate deposition, curation, annotation, and integrated analysis of genomic data for the mitochondrial disease clinical and research communities, Molecular Genetics and Metabolism. 114 (2015) 388–396. 10.1016/j.ymgme.2014.11.016.

[22] K.J. Karczewski, L.C. Francioli, D.G. MacArthur, The mutational constraint spectrum quantified from variation in 141,456 humans, Yearbook of Paediatric Endocrinology. (2020). 10.1530/ey.17.14.3.

[23] S.T. Sherry, M. Ward, K. Sirotkin, dbSNP—Database for Single Nucleotide Polymorphisms and Other Classes of Minor Genetic Variation, Genome Research. 9 (1999) 677–679. 10.1101/gr.9.8.677.

[24] M.J. Landrum, J.M. Lee, M. Benson, G.R. Brown, C. Chao, S. Chitipiralla, B. Gu, J. Hart, D. Hoffman, W. Jang, K. Karapetyan, K. Katz, C. Liu, Z. Maddipatla, A. Malheiro, K. McDaniel, M. Ovetsky, G. Riley, G. Zhou, J. Holmes, ClinVar: improving access to variant interpretations and supporting evidence, Nucleic Acids Research. 46 (2017) D1062–D1067. 10.1093/nar/gkx1153.

[25] Hedvig Tordai, O. Torres, Máté Csepi R. Padányi, G.L. Lukács, Tamás Hegedűs, Analysis of AlphaMissense data in different protein groups and structural context, Scientific Data. 11 (2024). 10.1038/s41597-024-03327-8.

[26] M. Mirdita, K. Schütze, Y. Moriwaki, L. Heo, S. Ovchinnikov, M. Steinegger, ColabFold: making protein folding accessible to all, Nature Methods. 19 (2022) 1–4. 10.1038/s41592-022-01488-1.

[27] M. Váradi, D. Bertoni, P. Magaña, U. Paramval, I. Pidruchna, M. Radhakrishnan, M. Tsenkov, S. Nair, M. Mirdita, J. Yeo, O. Kovalevskiy, K. Tunyasuvunakool, A. Laydon, A. Žídek, H. Tomlinson, D. Hariharan, J. Abrahamson, T. Green, J. Jumper, E. Birney, AlphaFold Protein Structure Database in 2024: providing structure coverage for over 214 million protein sequences, Nucleic Acids Research. 52 (2023). 10.1093/nar/gkad1011.

[28] A. Bateman, M.-J. Martin, S. Orchard, M. Magrane, A. Adesina, S. Ahmad, E.H. Bowler-Barnett, H. Bye-A-Jee, D. Carpentier, P. Denny, J. Fan, P. Garmiri, L. Jose, A. Hussein, A. Ignatchenko, G. Insana, R. Ishtiaq, V. Joshi, D. Jyothi, S. Kandasaamy, UniProt: the Universal Protein Knowledgebase in 2025, Nucleic Acids Research. 53 (2024). 10.1093/nar/gkae1010.

[29] D.M. Kruger, A. Ahmed, H. Gohlke, NMSim Web Server: integrated approach for normal mode-based geometric simulations of biologically relevant conformational transitions in proteins, Nucleic Acids Research. 40 (2012) W310–W316. 10.1093/nar/gks478.

[30] W. Humphrey, A. Dalke, K. Schulten, VMD: Visual molecular dynamics, Journal of Molecular Graphics. 14 (1996) 33–38. 10.1016/0263-7855(96)00018-5.

[31] A. Del Conte, G.F. Camagni, D. Clementel, G. Minervini, A.M. Monzon, C. Ferrari, D. Piovesan, S.E. Tosatto, RING 4.0: faster residue interaction networks with novel interaction types across over 35,000 different chemical structures, Nucleic Acids Research. (2024). 10.1093/nar/gkae337.

[32] R.C. Team, R: A language and environment for statistical computing., MSOR Connections. 1 (2014).

[33] RStudio Team, RStudio | Open Source & Professional Software for Data Science Teams, Rstudio.com. (2020). http://www.rstudio.com/.

[34] H. Wickham, Create Elegant Data Visualisations Using the Grammar of Graphics, Tidyverse.org. (2020). https://ggplot2.tidyverse.org..

[35] A. Kassambara, rstatix: Pipe-Friendly Framework for Basic Statistical Tests, CRAN: Contributed Packages. (2019). 10.32614/cran.package.rstatix.

[36] A. Lex, N. Gehlenborg, H. Strobelt, R. Vuillemot, H. Pfister, UpSet: Visualization of Intersecting Sets, IEEE Transactions on Visualization and Computer Graphics. 20 (2014) 1983–1992. 10.1109/tvcg.2014.2346248.

[37] A. Kassambara, ggplot2 Based Publication Ready Plots, Rpkgs.datanovia.com. (2023). https://rpkgs.datanovia.com/ggpubr/.

[38] A. Kassambara, ggcorrplot: Visualization of a Correlation Matrix using “ggplot2,” R-Packages. (2019). https://cran.r-project.org/package=ggcorrplot x(accessed February 14, 2022).

[39] M.C. Russillo, V. Andreozzi, R. Erro, M. Picillo, M. Amboni, S. Cuoco, P. Barone, M.T. Pellecchia, Sex Differences in Parkinson’s Disease: From Bench to Bedside, Brain Sciences. 12 (2022) 917. 10.3390/brainsci12070917.

[40] P. Martinelli, M. Contin, C. Scaglione, R. Riva, F. Albani, A. Baruzzi, Levodopa pharmacokinetics and dyskinesias: are there sex-related differences?, Neurological Sciences. 24 (2003) 192–193. 10.1007/s10072-003-0125-z.

[41] H. Iwaki, C. Blauwendraat, H.L. Leonard, M.B. Makarious, J.J. Kim, G. Liu, J. Maple□ Grødem, J. Corvol, L. Pihlstrøm, M. Nimwegen, L. Smolensky, N. Amondikar, S.J. Hutten, M. Frasier, K.H. Nguyen, J. Rick, S. Eberly, F. Faghri, P. Auinger, K.M. Scott, Differences in the Presentation and Progression of Parkinson’s Disease by Sex, Movement Disorders. (2020). 10.1002/mds.28312.

[42] M. Tió-Coma, S.M. Kiełbasa, S. J.F, H. Mei, Johan Chandra Roy, J. Wallinga, M. Khatun, S. Soren, Abu Sufian Chowdhury, K. Alam, Anouk van Hooij, Jan Hendrik Richardus, Annemieke Geluk, Blood RNA signature RISK4LEP predicts leprosy years before clinical onset, EBioMedicine (Amsterdam). 68 (2021) 103379–103379. 10.1016/j.ebiom.2021.103379.

[43] C. Wirth, U. Brandt, C. Hunte, V. Zickermann, Structure and function of mitochondrial complex I, Biochimica et Biophysica Acta (BBA) - Bioenergetics. 1857 (2016) 902–914. 10.1016/j.bbabio.2016.02.013.

[44] V.K. Moparthi, B. Kumar, Y. Al-Eryani, E. Sperling, K. Górecki, T. Drakenberg, C. Hägerhäll, Functional role of the MrpA- and MrpD-homologous protein subunits in enzyme complexes evolutionary related to respiratory chain complex I, Biochimica et Biophysica Acta (BBA) - Bioenergetics. 1837 (2013) 178–185. 10.1016/j.bbabio.2013.09.012.

[45] V. Zickermann, C. Wirth, H. Nasiri, K. Siegmund, H. Schwalbe, C. Hunte, U. Brandt, Mechanistic insight from the crystal structure of mitochondrial complex I, Science. 347 (2015) 44–49. 10.1126/science.1259859.

[46] L. Euro, G. Belevich, M.I. Verkhovsky, M. Wikström, M. Verkhovskaya, Conserved lysine residues of the membrane subunit NuoM are involved in energy conversion by the proton-pumping NADH:ubiquinone oxidoreductase (Complex I), Biochimica et Biophysica Acta. Bioenergetics. 1777 (2008) 1166–1172. 10.1016/j.bbabio.2008.06.001.

[47] J. Torres-Bacete, P.K. Sinha, A. Matsuno-Yagi, T. Yagi, Structural Contribution of C-terminal Segments of NuoL (ND5) and NuoM (ND4) Subunits of Complex I from Escherichia coli, Journal of Biological Chemistry. 286 (2011) 34007–34014. 10.1074/jbc.m111.260968.

[48] E. Mkaouar-Rebai, M. Ammar, L. Sfaihi, O. Alila-Fersi, M. Maalej, R. Felhi, M. Hachicha, F. Fakhfakh, Mitochondrial disease patients with novel ND4 12058A > C and ND1 m.3911A > G variations: implications for a role in the phenotype following a bioinformatic investigation, Molecular Biology Reports. (2021). 10.1007/s11033-021-06452-4.

[49] S. Ellouze, S. Augustin, A. Bouaita, C. Bonnet, M. Simonutti, V. Forster, S. Picaud, J.-A. Sahel, M. Corral-Debrinski, Optimized Allotopic Expression of the Human Mitochondrial ND4 Prevents Blindness in a Rat Model of Mitochondrial Dysfunction, The American Journal of Human Genetics. 83 (2008) 373–387. 10.1016/j.ajhg.2008.08.013.

[50] W. Yan, Y. Kang, X. Ji, S. Li, Y. Li, G. Zhang, H. Cui, G. Shi, Testosterone Upregulates the Expression of Mitochondrial ND1 and ND4 and Alleviates the Oxidative Damage to the Nigrostriatal Dopaminergic System in Orchiectomized Rats, Oxidative Medicine and Cellular Longevity. 2017 (2017) 1–13. 10.1155/2017/1202459.

[51] D. Pignataro, S. Francia, F. Zanetta, G. Brenna, S. Brandini, A. Olivieri, A. Torroni, G. Biamonti, A. Montecucco, A missense MT-ND5 mutation in differentiated Parkinson Disease cytoplasmic hybrid induces ROS-dependent DNA Damage Response amplified by DROSHA, Scientific Reports. 7 (2017). 10.1038/s41598-017-09910-x.

[52] T. Danhelovska, H. Kolarova, J. Zeman, H. Hansikova, M. Vaneckova, L. Lambert, V. Kucerova-Vidrova, K. Berankova, T. Honzik, M. Tesarova, Multisystem mitochondrial diseases due to mutations in mtDNA-encoded subunits of complex I, BMC Pediatrics. 20 (2020). 10.1186/s12887-020-1912-x.

[53] H. Fang, H. Shi, X. Li, D. Sun, F. Li, B. Li, Y. Ding, Y. Ma, Y. Liu, Y. Zhang, L. Shen, Y. Bai, Y. Yang, J. Lu, Exercise intolerance and developmental delay associated with a novel mitochondrial ND5 mutation, Scientific Reports. 5 (2015). 10.1038/srep10480.

[54] M.L. Valentino, P. Barboni, C. Rengo, A. Achilli, A. Torroni, R. Lodi, C. Tonon, B. Barbiroli, F. Fortuna, P. Montagna, A. Baruzzi, V. Carelli, The 13042G->A/ND5 mutation in mtDNA is pathogenic and can be associated also with a prevalent ocular phenotype, Journal of Medical Genetics. 43 (2005) e38–e38. 10.1136/jmg.2005.037507.

