## Supplementary Material for "MOLECULAR ANALYSIS OF MITOCHONDRIAL COMPLEX I IN THE RESPONSIVENESS TO LEVODOPA IN PARKINSON’S DISEASE"

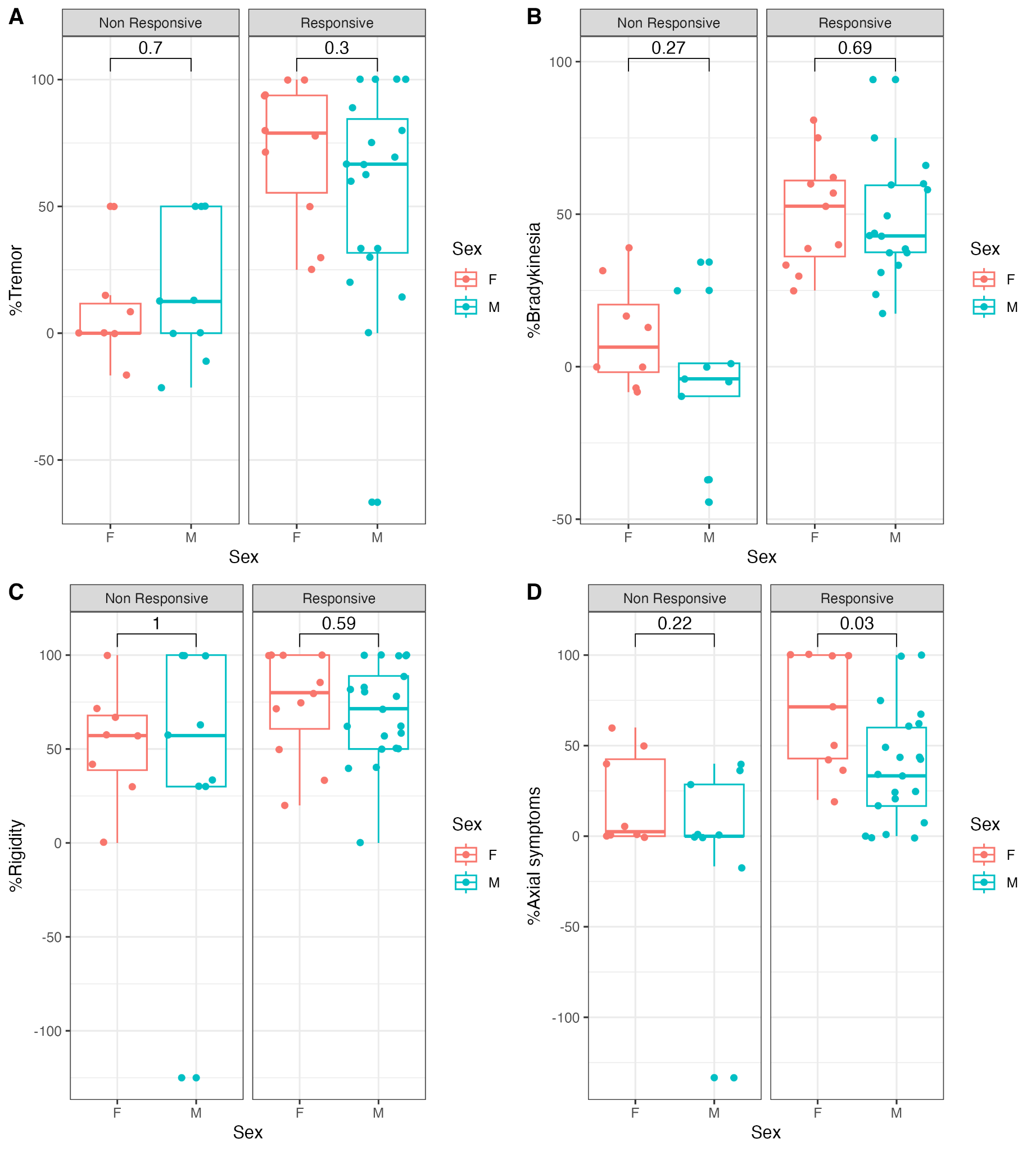

**Figure S1:** Motor characteristics in response to levodopa according to sex by responsiveness group. A: Percentage change in tremor between sexes and responsiveness to treatment, B: Percentage change in bradykinesia between sexes and responsiveness to treatment, C: Percentage change in rigidity between sexes and responsiveness to treatment, D: Percentage change in axial symptoms between sexes and responsiveness to treatment. F: female; M: male

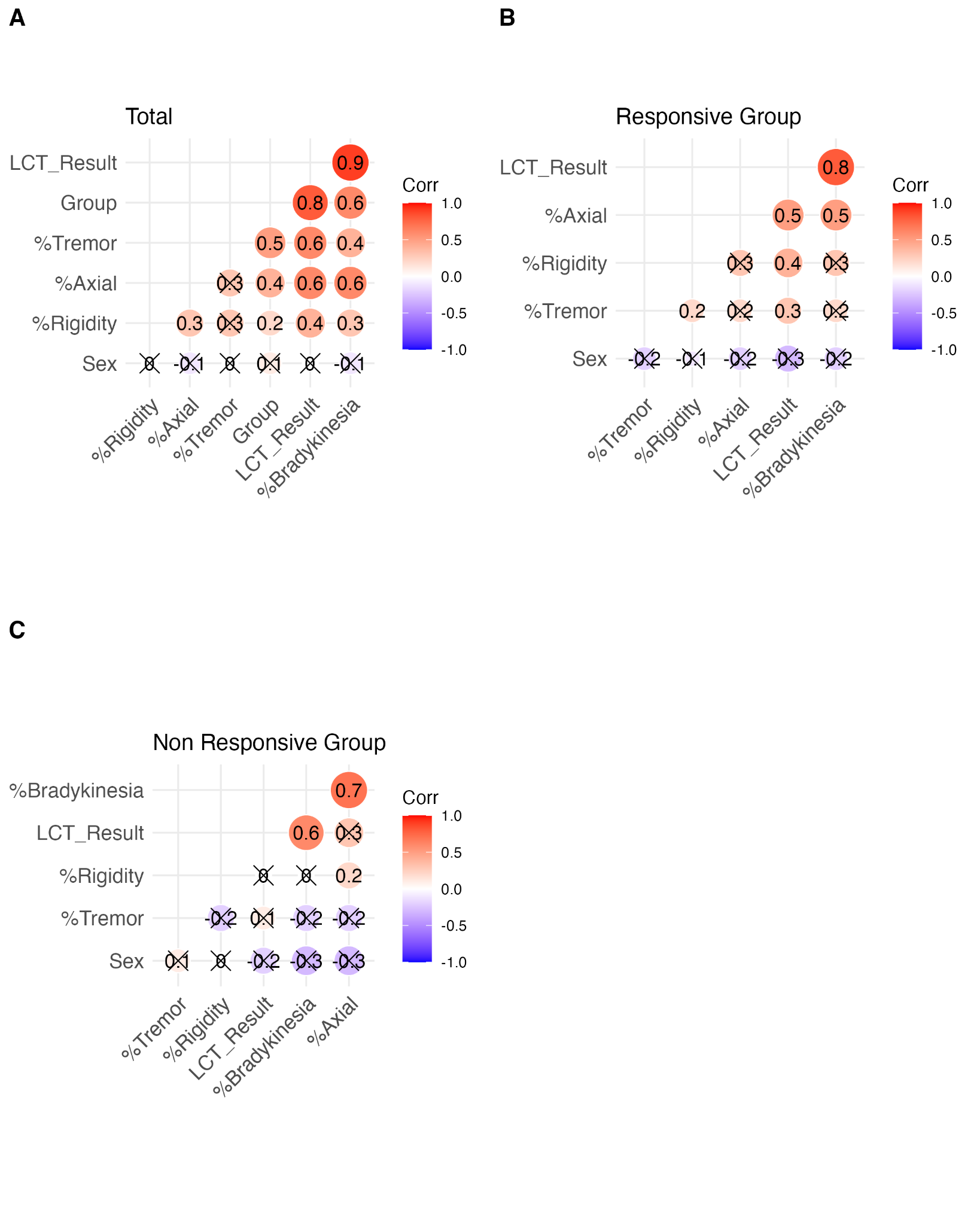

**Figure S2:** Spearman correlation analysis between motor characteristics related to response to levodopa. A: Correlation between motor characteristics of the total cohort. B: Correlation between motor characteristics of the LCT-responsive group. C: Correlation between motor characteristics of the LCT-nonresponsive group. The colors of the figures indicate significant p-values, with those closest to red indicating positive correlations and those closest to blue indicating negative correlations, while circles marked with X are nonsignificant p-values (P>0.05).

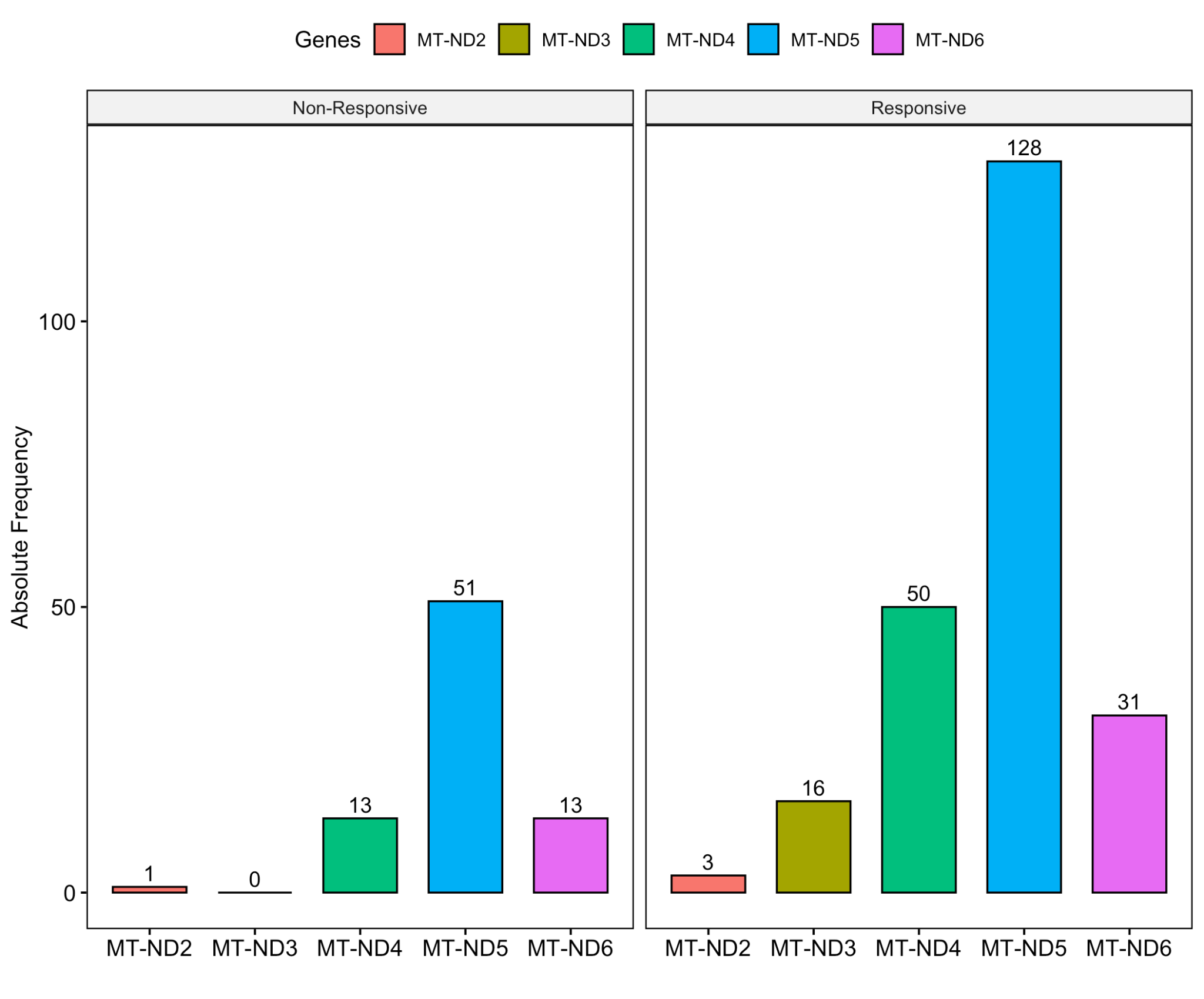

**Figure S3.** Distribution of unique heteroplasmic variants per gene according to non-responsive and responsive groups to LCT.

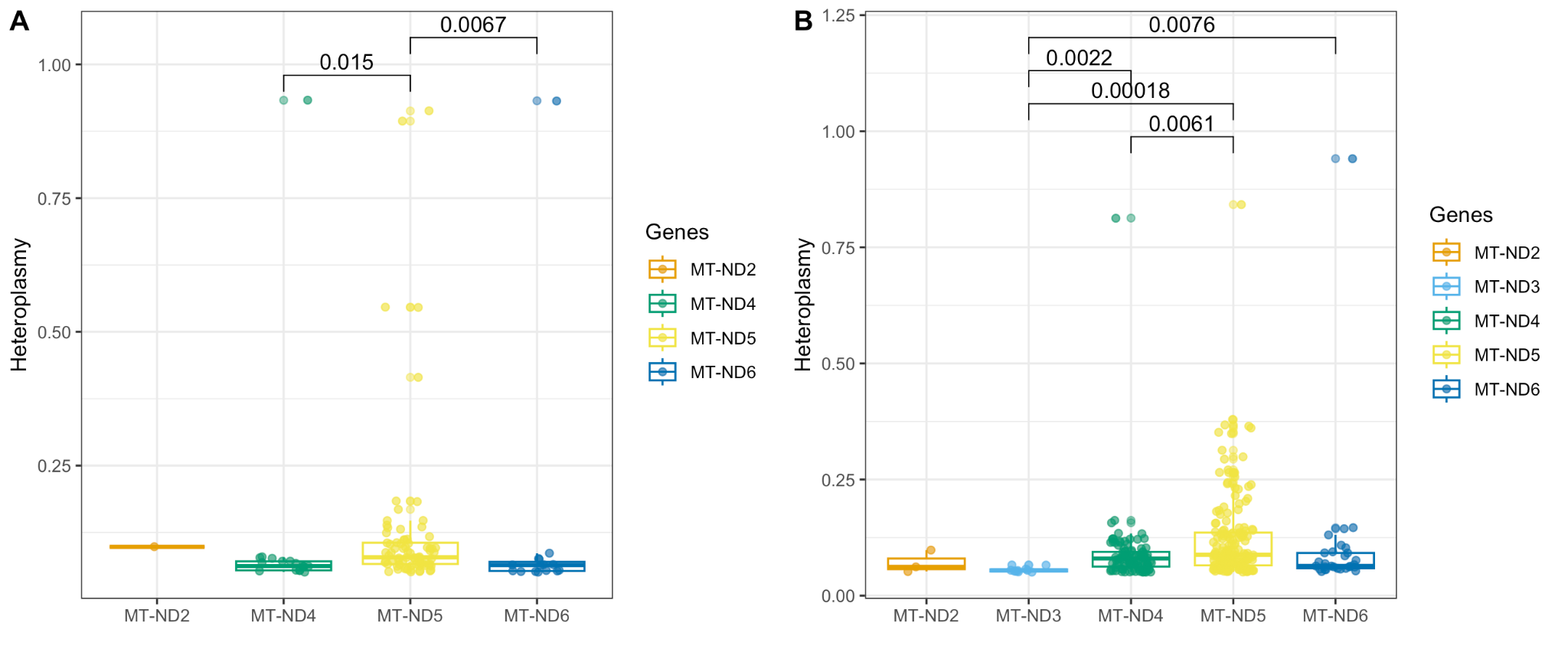

**Figure S4.** Heteroplasmy of variants of mitochondrial genes encoding subunits of complex I. A: Distribution of heteroplasmy of variants from patients unresponsive to LCT. B: Distribution of heteroplasmy of variants from patients responsive to LCT.

**Table S1.** Most frequent mutations in patients responsive to LCT. Most frequent mutations (present in at least 60% of the cohort) of the CI of patients with LCT responsiveness and their clinical, genomic and proteomic consequences already described in databases.

| **Gene** | **Mutation** | **Consequence** | **dbSNP** | **Polyphen** | **SIFT** | **AlphaMissense (score)** | **ClinVar** | **Condition (ClinVar)** |
| --- | --- | --- | --- | --- | --- | --- | --- | --- |
| *MT-ND4* | 11505T>C | Missense | . | Benign | Tolerable - low confidence | Ambiguous (0.441) | . | . |
|  | 11963G>A | Missense | rs201803948 | Benign | Tolerable - low confidence | Probably benign (0.077) | Benign | Leigh syndrome |
|  | 12018C>G | Missense | . | . | . | Probably benign (0.11) | . | . |
|  | 12013A>G | Synonym | rs1057516067 | . | . | . | Uncertain significance | Extrapyramidal basal ganglia calcification, Dementia, Developmental delay, Encephalopathy, Epilepsy, Hypotonia, Congenital cardiomyopathy, Developmental delay, Hearing impairment, Microcephaly, Developmental delay, Hearing impairment, Macrocephaly, Leigh syndrome |
|  | 11788C>T | Synonym | rs1556423994 | . | . | . | . | . |
|  | 11887G>A | Synonym | rs1556424007 | . | . | . | . | . |
|  | 11770T>C | Synonym | rs1603223408 | . | . | . | . | . |
|  | 11809T>C | Synonym | rs1603223420 | . | . | . | . | . |
|  | 11827T>C | Synonym | rs368026942 | . | . | . | . | . |
| *MT-ND5* | 13130C>A | Missense | . | . | . | Ambiguous (0.342) | . | . |
|  | 13130C>T | Missense | . | . | . | Ambiguous (0.497) | . | . |
|  | 13845C>T | Synonym | . | . | . | . | . | . |
|  | 13174T>C | Synonym | rs1556424231 | . | . | . | . | . |
|  | 13359G>A | Synonym | rs1556424268 | . | . | . | . | . |
|  | 13260T>C | Synonym | rs1603224109 | . | . | . | . | . |
|  | 13317G>A | Synonym | rs1603224142 | . | . | . | . | . |
|  | 13386T>C | Synonym | rs1603224171 | . | . | . | . | . |
|  | 13581T>C | Synonym | rs1603224247 | . | . | . | . | . |
|  | 13401T>C | Synonym | rs28379170 | . | . | . | . | . |
|  | 13368G>A | Synonym | rs3899498 | . | . | . | . | . |
|  | 13488T>C | Synonym | rs878853050 | . | . | . | Probably benign | . |

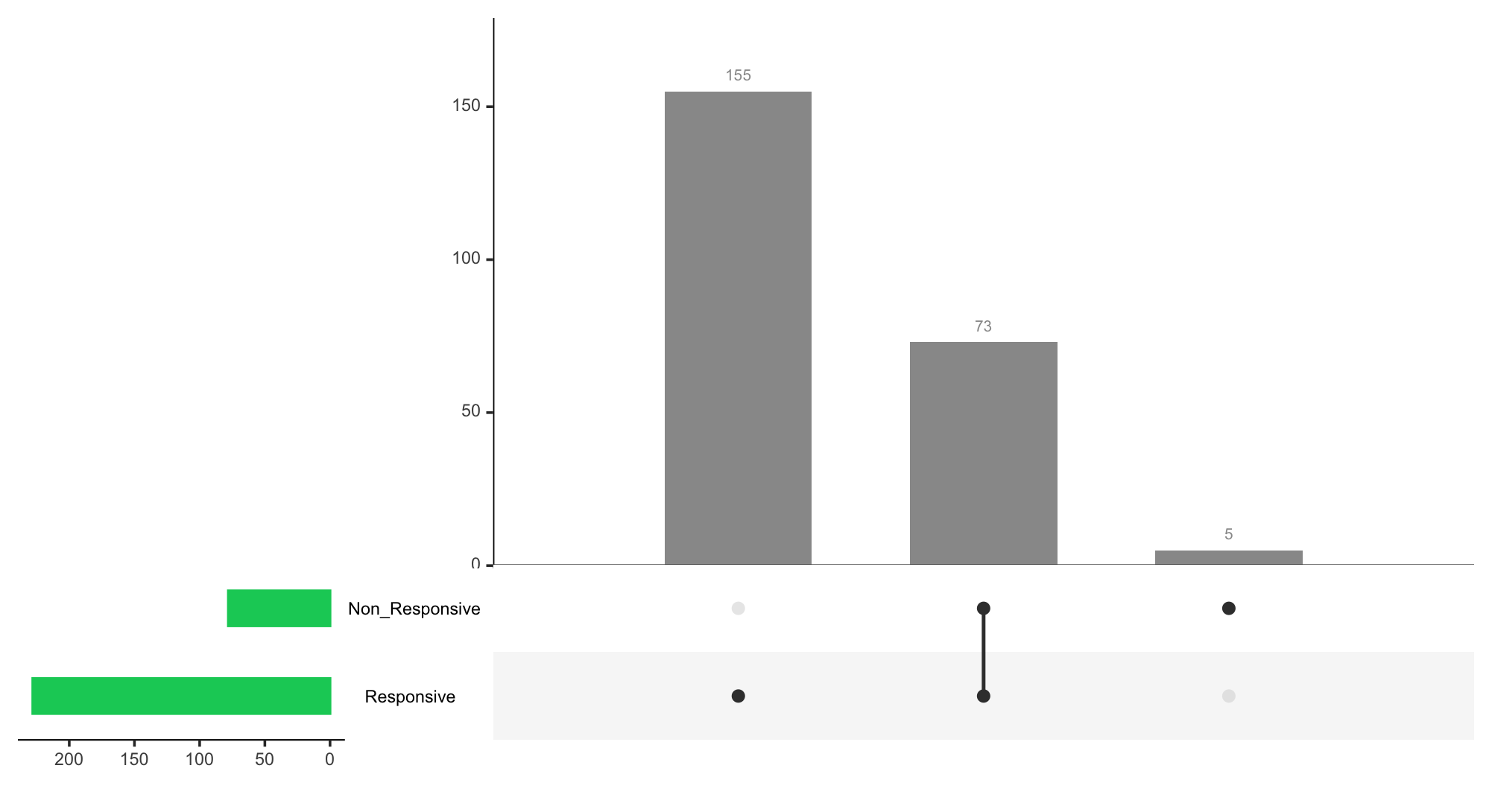

**Figure S5.** Distribution of mitochondrial variants found in the LCT-responsive and non-responsive groups. Each dark dot indicates the group with the respective number of variants (gray columns), and the line between dots represents the intersection between the groups. The set size (green columns) is the overall number of variants.

**Tabela S2.** Mutations exclusive to the non-responsive group in LCT. Mutations of the CI exclusive to patients with low responsiveness in LCT and their clinical, genomic and proteomic consequences already described in databases.

| **Gene** | **Mutation** | **Heteroplasmy** | **Consequence** | **dbSNP** | **Polyphen** | **SIFT** | **AlphaMissense (score)** | **ClinVar** | **Condition (ClinVar)** |
| --- | --- | --- | --- | --- | --- | --- | --- | --- | --- |
| *MT-ND2* | 5322A>C | 0.098 | Missense | . | Benign | Tolerable | Probably benign (0.096) | . | . |
| *MT-ND4* | 12010C>T | 0.933 | Synonym | rs1057516066 | . | . | . | Uncertain significance | Dystonic disorder, developmental delay |
| *MT-ND5* | 13989C>T | 0.913 | Synonym | rs1603224457 | . | . | . | . | . |
|  | 12612A>G | 0.894 | Synonym | rs28359172 | . | . | . | . | . |
| *MT-ND6* | 14170A>T | 0.052 | Missense | . | . | . | Probably benign (0.107) | . | . |

**Table S3.** Most frequent unique mutations in LCT-responsive patients. Most frequent unique mutations (present in at least 60% of the cohort) of the CI of patients with LCT responsiveness and their clinical, genomic and proteomic consequences already described in databases.

| **Gene** | **Mutation** | **Consequence** | **dbSNP** | **Polyphen** | **SIFT** | **AlphaMissense (score)** | **ClinVar** | **Condition (ClinVar)** |
| --- | --- | --- | --- | --- | --- | --- | --- | --- |
| *MT-ND4* | 11827T>C | Synonym | rs3899498 | . | . | . | . | . |
| *MT-ND5* | 13368G>A | Synonym | rs368026942 | . | . | . | . | . |
|  | 13317G>A | Synonym | rs1603224142 | . | . | . | . | . |
|  | 13401T>C | Synonym | rs28379170 | . | . | . | . | . |

**Table S4.** Mutations shared by the non-responsive and responsive groups in LCT. CI mutations present in both groups of patients in relation to LCT responsiveness and their clinical, genomic and proteomic consequences already described in databases.

| **Gene** | **Mutation** | **Consequence** | **dbSNP** | **Polyphen** | **SIFT** | **AlphaMissense (score)** | **ClinVar** | **Condition (ClinVar)** |
| --- | --- | --- | --- | --- | --- | --- | --- | --- |
| *MT-ND4* | 11600G>A | Missense | . | Probably damaging | Deleterious - low confidence | Probably pathogenic (0.91) | . | . |
|  | 12018C>G | Missense | . | . | . | Probably benign (0.11) | . | . |
|  | 11505T>C | Missense | . | Benign | Tolerable - low confidence | Ambígua (0.441) | . | . |
|  | 11963G>A | Missense | rs201803948 | Benign | Tolerable - low confidence | Ambiguous (0.077) | Benign | Leigh syndrome |
|  | 11914G>A | Synonym | rs2853496 | . | . | . | . | . |
|  | 12007G>A | Synonym | rs2853497 | . | . | . | Benign | Venous thromboembolism |
|  | 12112C>T | Synonym | rs28695839 | . | . | . | . | . |
|  | 11770T>C | Synonym | rs1603223408 | . | . | . | . | . |
|  | 11809T>C | Synonym | rs1603223420 | . | . | . | . | . |
|  | 11788C>T | Synonym | rs1556423994 | . | . | . | . | . |
|  | 11887G>A | Synonym | rs1556424007 | . | . | . | . | . |
|  | 12013A>G | Synonym | rs1057516067 | . | . | . | Uncertain significance | Extrapyramidal basal ganglia calcification, Dementia, Developmental delay, Encephalopathy, Epilepsy, Hypotonia, Congenital cardiomyopathy, Developmental delay, Hearing impairment, Microcephaly, Developmental delay, Hearing impairment, Macrocephaly, Leigh syndrome |
| *MT-ND5* | 13902C>T | Synonym | . | . | . | . | . | . |
|  | 14016G>A | Synonym | rs1556424367 | . | . | . | . | . |
|  | 14088T>C | Synonym | rs193302974 | . | . | . | . | . |
|  | 14034T>C | Synonym | rs60673542 | . | . | . | . | . |
|  | 14097C>T | Synonym | rs1556424382 | . | . | . | . | . |
|  | 14122A>C | Missense | . | Benign | Tolerable - low confidence | Probably benign (0.058) | . | . |
|  | 13635T>C | Synonym | rs878853075 | . | . | . | Likely benign | Not provided |
|  | 13644C>T | Synonym | . | . | . | . | . | . |
|  | 13813G>A | Missense | rs1556424332 | Benign | Tolerable - low confidence | Probably benign (0.099) | Benign | Leigh syndrome |
|  | 13833A>G | Synonym | rs1603224376 | . | . | . | . | . |
|  | 14040G>A | Synonym | rs57180882 | . | . | . | Probably benign | Not provided |
|  | 14053A>G | Missense | rs200134839 | Benign | Tolerable - low confidence | Probably benign (0.068) | Benign | Leigh syndrome |
|  | 13028C>T | Missense | . | . | . | Probably pathogenic (0.8) | . | . |
|  | 13638A>G | Synonym | rs1603224278 | . | . | . | . | . |
|  | 13650C>A | Synonym | . | . | . | . | . | . |
|  | 13811C>G | Missense | . | . | . | Probably benign (0.132) | . | . |
|  | 13879T>A | Missense | rs879087566 | Benign | Tolerable - low confidence | Probably benign (0.074) | Probably benign | Leigh syndrome |
|  | 13929C>T | Synonym | . | . | . | . | . | . |
|  | 13950C>T | Synonym | . | . | . | . | . | . |
|  | 13980G>A | Synonym | rs1569484618 | . | . | . | . | . |
|  | 14001A>G | Synonym | rs1603224465 | . | . | . | . | . |
|  | 14002A>G | Missense | rs386829198 | Benign | Tolerable - low confidence | Probably benign (0.101) | Benign/Probably benign | Not provided, Leigh syndrome |
|  | 14020T>C | Synonym | rs1556424369 | . | . | . | . | . |
|  | 13111T>C | Synonym | rs1556424221 | . | . | . | . | . |
|  | 13164A>C | Synonym |  | . | . | . | . | . |
|  | 13812T>C | Synonym | rs1556424331 | . | . | . | . | . |
|  | 13820T>C | Missense | rs1603224368 | Benign | Tolerable - low confidence | Probably benign (0.172) | Benign | Leigh syndrome |
|  | 13890C>T | Synonym | rs1603224404 | . | . | . | . | . |
|  | 13968G>A | Synonym | rs1603224439 | . | . | . | . | . |
|  | 13281T>C | Synonym | rs879154628 | . | . | . | . | . |
|  | 13809C>A | Synonym | . | . | . | . | . | . |
|  | 13869T>C | Synonym | rs1556424339 | . | . | . | . | . |
|  | 13887A>G | Synonym | rs1603224401 | . | . | . | . | . |
|  | 13889G>A | Missense | rs1556424343 | Benign | Tolerable - low confidence | Probably benign (0.103) | Benign | Leigh syndrome |
|  | 13899T>C | Synonym | rs370031192 | . | . | . | . | . |
|  | 13905C>T | Synonym | . | . | . | . | . | . |
|  | 13908C>T | Synonym | . | . | . | . | . | . |
|  | 13920C>T | Synonym | . | . | . | . | . | . |
|  | 13934C>T | Missense | rs193302971 | Benign | Tolerable - low confidence | Probably benign (0.073) | Benign | Leigh syndrome |
|  | 13945A>G | Missense | rs1603224426 | Benign | Tolerable - low confidence | Probably benign (0.089) | . | . |
|  | 13260T>C | Synonym | rs1603224109 | . | . | . | . | . |
|  | 13359G>A | Synonym | rs1556424268 | . | . | . | . | . |
|  | 13386T>C | Synonym | rs1603224171 | . | . | . | . | . |
|  | 13845C>T | Synonym | . | . | . | . | . | . |
|  | 13130C>A | Missense | . | . | . | Ambiguous (0.342) | . | . |
|  | 13130C>T | Missense | . | . | . | Ambiguous (0.497) | . | . |
|  | 13174T>C | Synonym | rs1556424231 | . | . | . | . | . |
|  | 13488T>C | Synonym | rs878853050 | . | . | . | . | . |
|  | 13581T>C | Synonym | rs1603224247 | . | . | . | . | . |
| *MT-ND6* | 14149C>T | Stop retained | rs1603224562 | . | . | . | . | . |
|  | 14311T>C | Synonym | rs1556424422 | . | . | . | . | . |
|  | 14155C>A | Synonym | . | . | . | . | . | . |
|  | 14215T>C | Synonym | rs879144433 | . | . | . | . | . |
|  | 14318T>C | Missense | rs28357675 | Benign | Tolerable | Probably benign (0.061) | Benign | Leigh syndrome |
|  | 14364G>A | Synonym | rs879086798 | . | . | . | . | . |
|  | 14368C>T | Synonym | . | . | . | . | . | . |
|  | 14182T>C | Synonym | rs372515139 | . | . | . | . | . |
|  | 14218T>C | Synonym | rs28553869 | . | . | . | . | . |
|  | 14305G>A | Synonym | rs386829208 | . | . | . | . | . |
|  | 14323G>A | Synonym | rs879208488 | . | . | . | . | . |
|  | 14362C>T | Synonym | . | . | . | . | . | . |

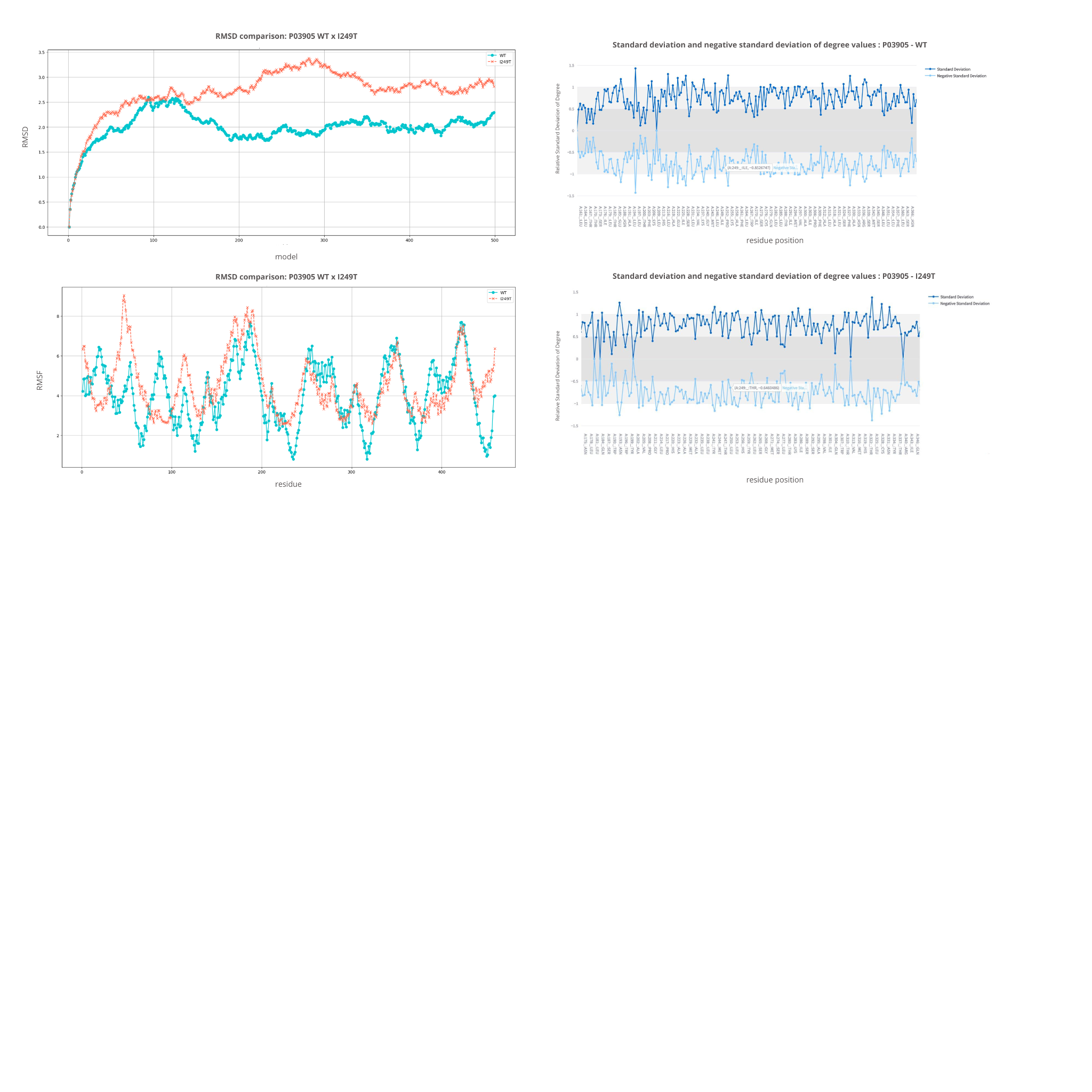

**Figure S6.** Structural comparison and analysis of dynamic variations of protein P03905, in wild-type (WT) and mutant (I249T) forms. A: Comparison of RMSD values of conformational variants along the simulated trajectory for WT (blue) and I249T (orange). B: Comparison of RMSF values per residue for WT (blue) and I249T (orange). C: Standard deviation and negative standard deviation of the number of bonds for P03905 (WT), evidencing variations in the stability of interactions observed in the conformational variants obtained in the dynamics. D: Standard deviation and negative standard deviation of the number of bonds for P03905 in the mutant form I249T, also demonstrating changes in the interaction patterns.

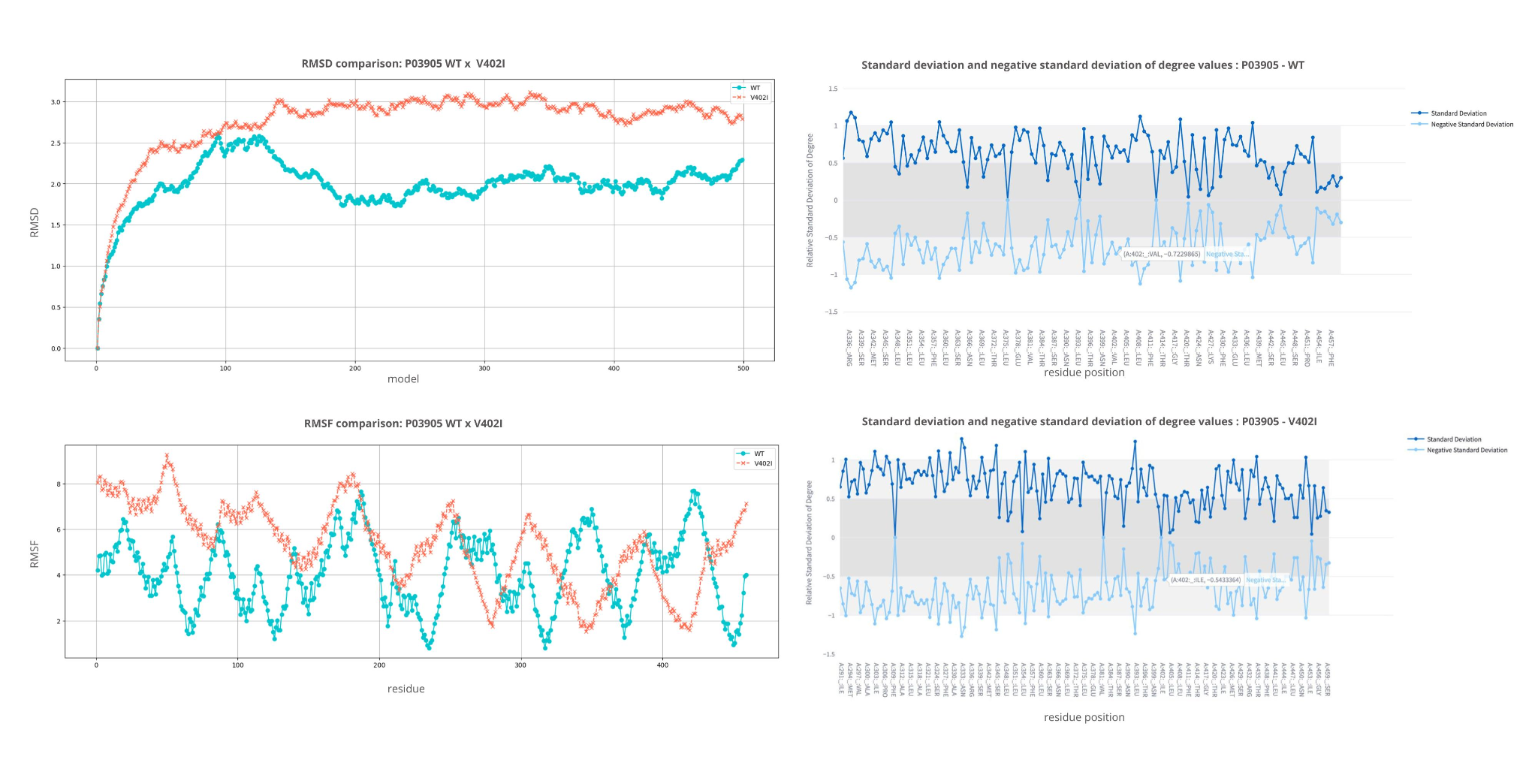

**Figure S7.** Structural comparison and analysis of dynamic variations of the P03905 protein, in the wild-type (WT) and mutant (V402I) forms. A: Comparison of RMSD values of the conformational variants along the simulated trajectory for WT (blue) and V402I (orange). B: Comparison of RMSF values per residue for WT (blue) and V402I (orange). C: Standard deviation and negative standard deviation of the number of bonds for P03905 (WT), evidencing variations in the stability of interactions observed in the conformational variants obtained in the dynamics. D: Standard deviation and negative standard deviation of the number of bonds for P03905 in the mutant form V402I, also demonstrating changes in the interaction patterns.

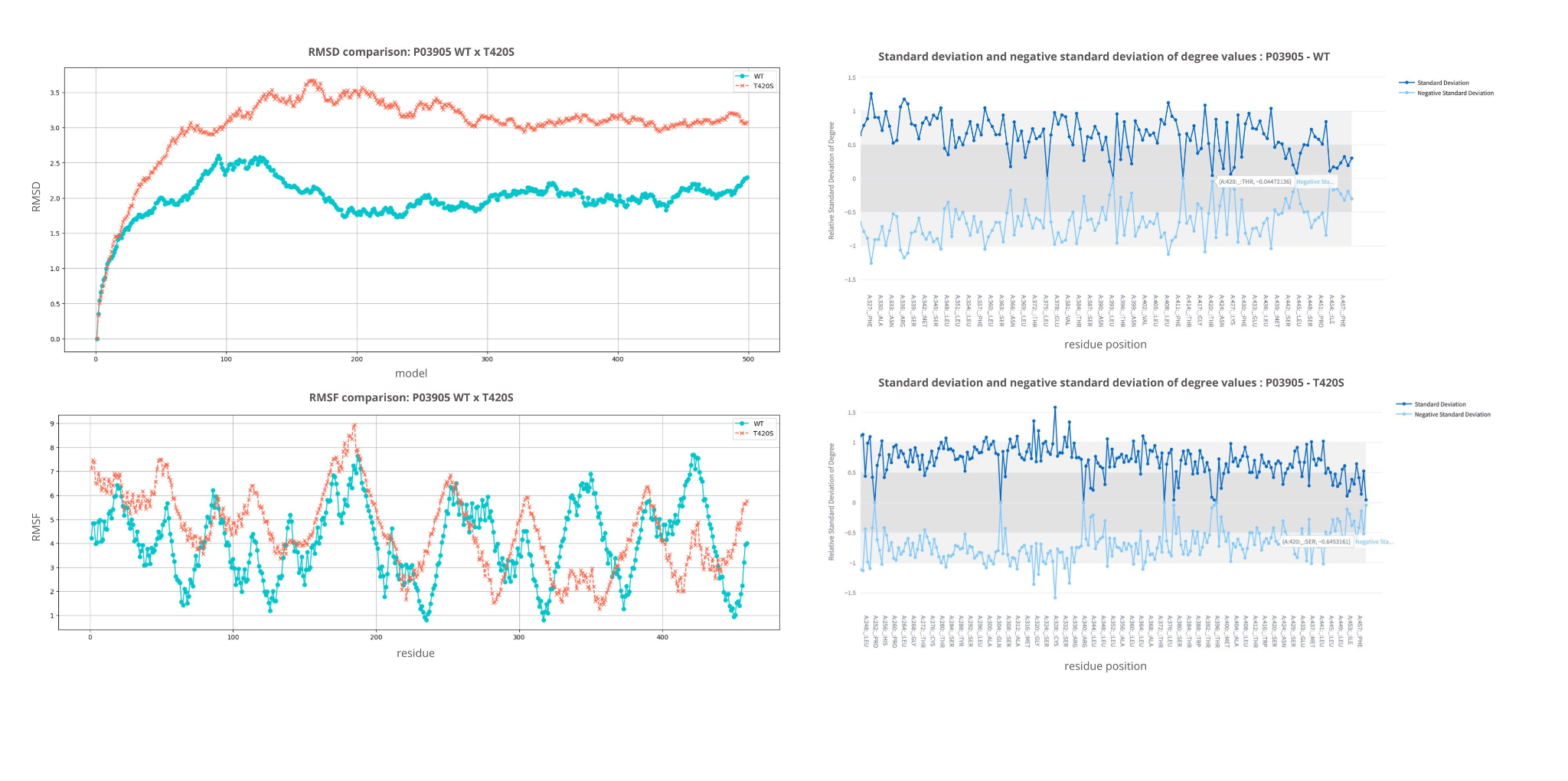
**Figure S8.** Structural comparison and analysis of dynamic variations of the P03905 protein, in the wild-type (WT) and mutant (T420S) forms. A: Comparison of the RMSD values of the conformational variants along the simulated trajectory for WT (blue) and T420S (orange). B: Comparison of the RMSF values per residue for WT (blue) and T420S (orange). C: Standard deviation and negative standard deviation of the number of bonds for P03905 (WT), evidencing variations in the stability of interactions observed in the conformational variants obtained in the dynamics. D: Standard deviation and negative standard deviation of the number of bonds for P03905 in the T420S mutant form, also demonstrating changes in the interaction patterns.

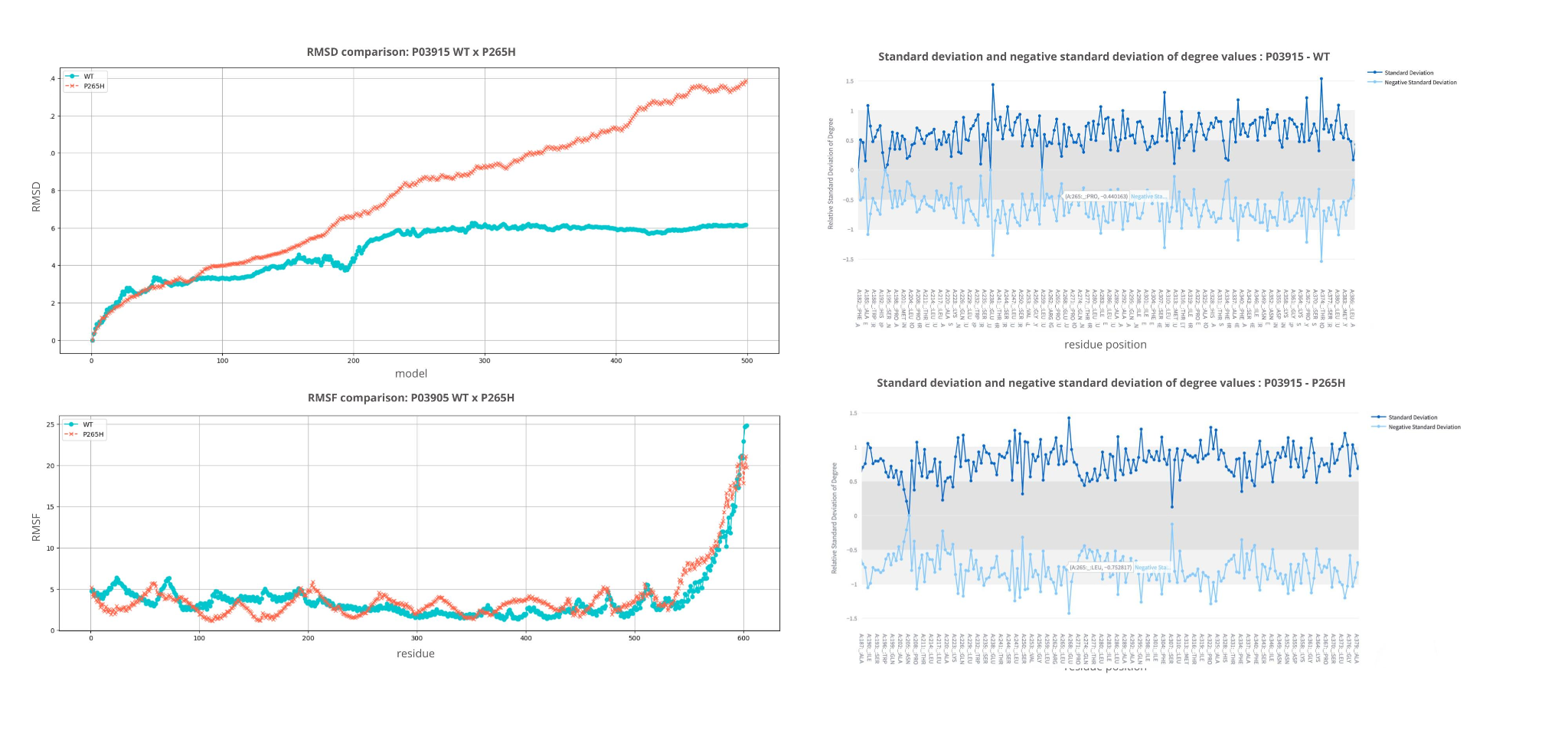

**Figure S9.** Structural comparison and analysis of dynamic variations of the P03915 protein, in the wild-type (WT) and mutant (P265H) forms. A: Comparison of RMSD values of the conformational variants along the simulated trajectory for WT (blue) and P265H (orange). B: Comparison of RMSF values per residue for WT (blue) and P265H (orange). C: Standard deviation and negative standard deviation of the number of bonds for P03915 (WT), evidencing variations in the stability of interactions observed in the conformational variants obtained in the dynamics. D: Standard deviation and negative standard deviation of the number of bonds for P03915 in the mutant form P265H, also demonstrating changes in the interaction patterns.

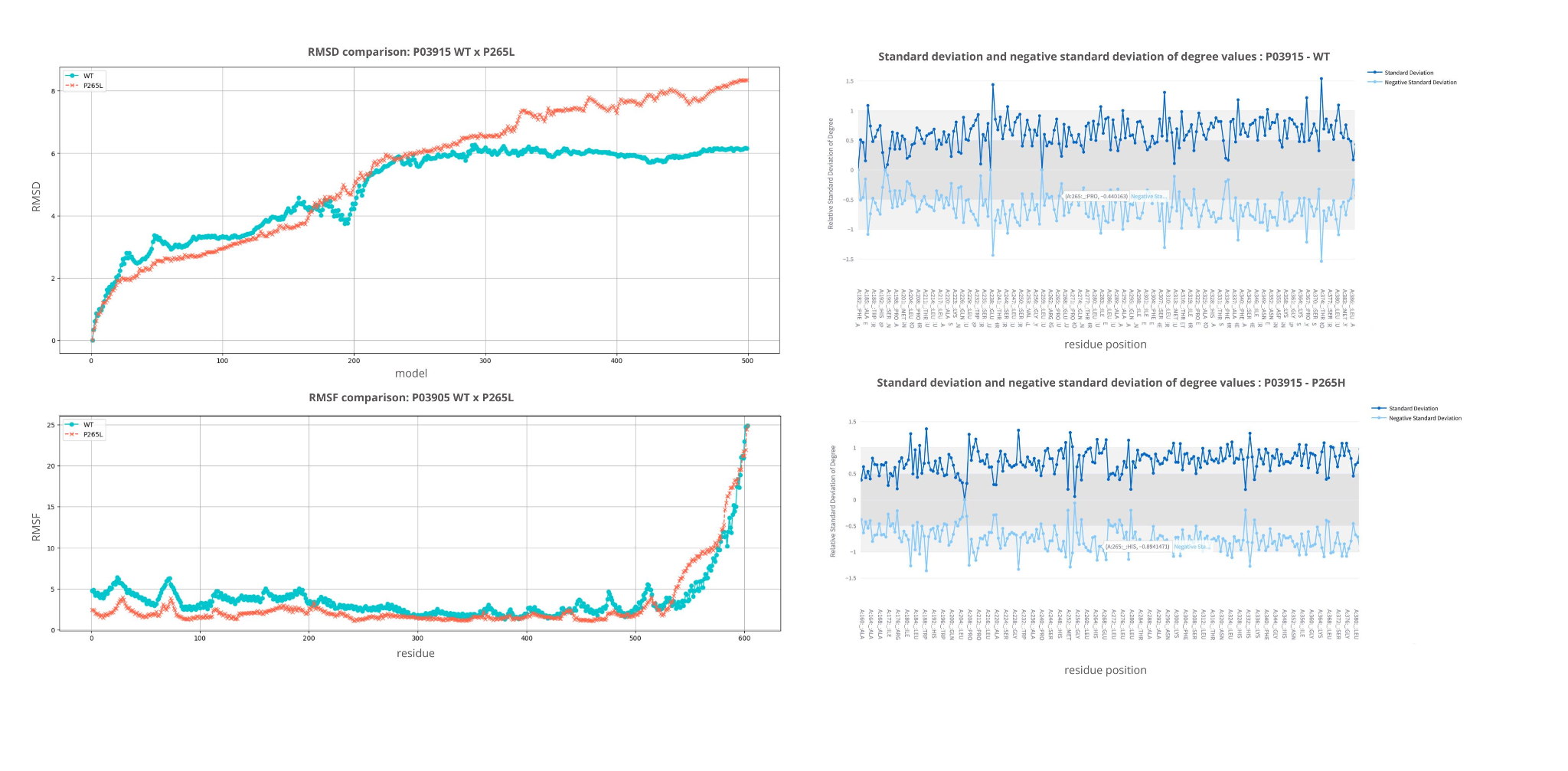
**Figure S10.** Structural comparison and analysis of dynamic variations of the P03915 protein, in the wild-type (WT) and mutant (P265L) forms. A: Comparison of RMSD values of the conformational variants along the simulated trajectory for WT (blue) and P265L (orange). B: Comparison of RMSF values per residue for WT (blue) and P265L (orange). C: Standard deviation and negative standard deviation of the number of bonds for P03915 (WT), evidencing variations in the stability of interactions observed in the conformational variants obtained in the dynamics. D: Standard deviation and negative standard deviation of the number of bonds for P03915 in the mutant form P265L, also demonstrating changes in the interaction patterns.

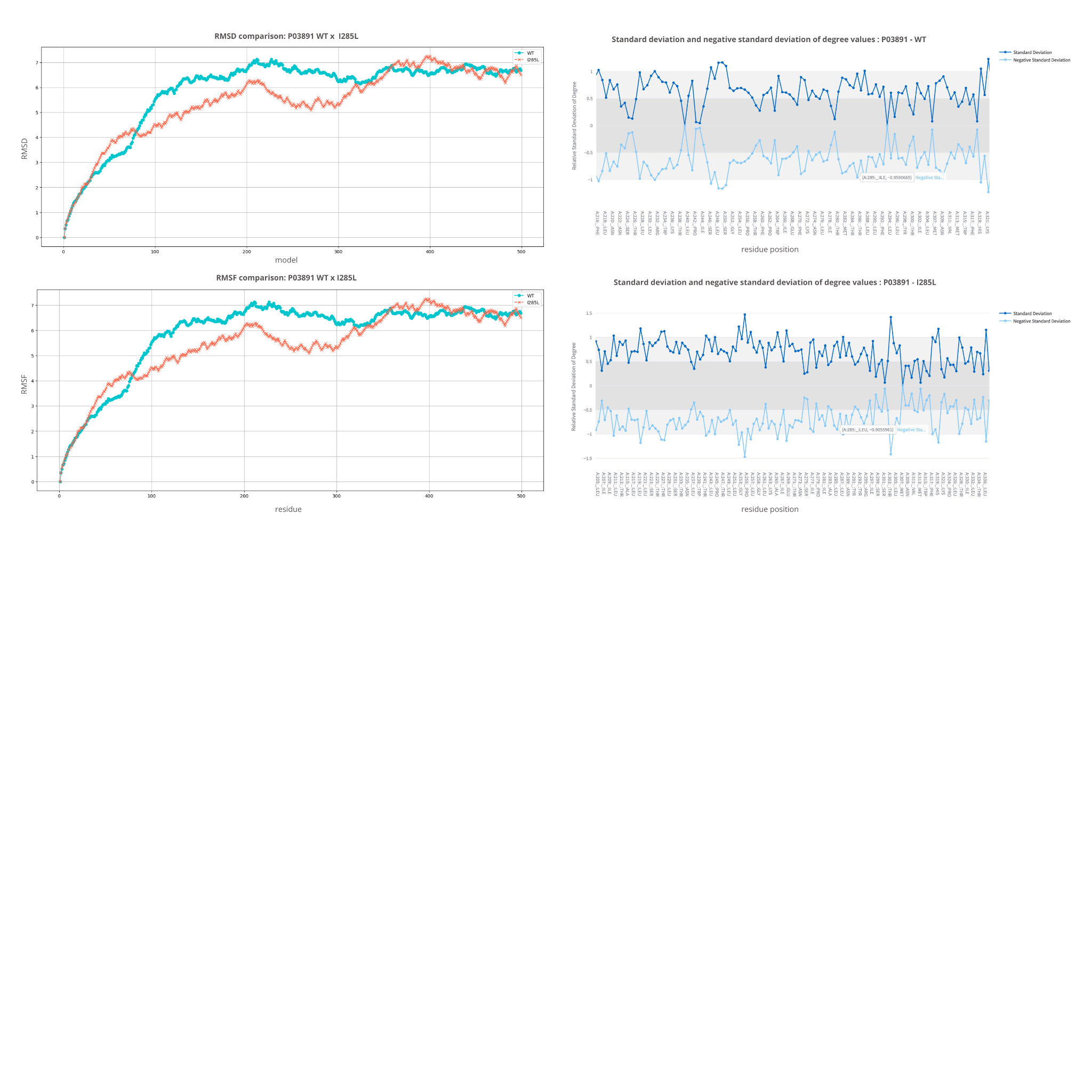

**Figure S11.** Structural comparison and analysis of dynamic variations of the P03891 protein, in the wild-type (WT) and mutant (I285L) forms. A: Comparison of RMSD values of the conformational variants along the simulated trajectory for WT (blue) and I285L (orange). B: Comparison of RMSF values per residue for WT (blue) and I285L (orange). C: Standard deviation and negative standard deviation of the number of bonds for P03891 (WT), evidencing variations in the stability of interactions observed in the conformational variants obtained in the dynamics. D: Standard deviation and negative standard deviation of the number of bonds for P03891 in the mutant form I285L, also demonstrating changes in the interaction patterns.

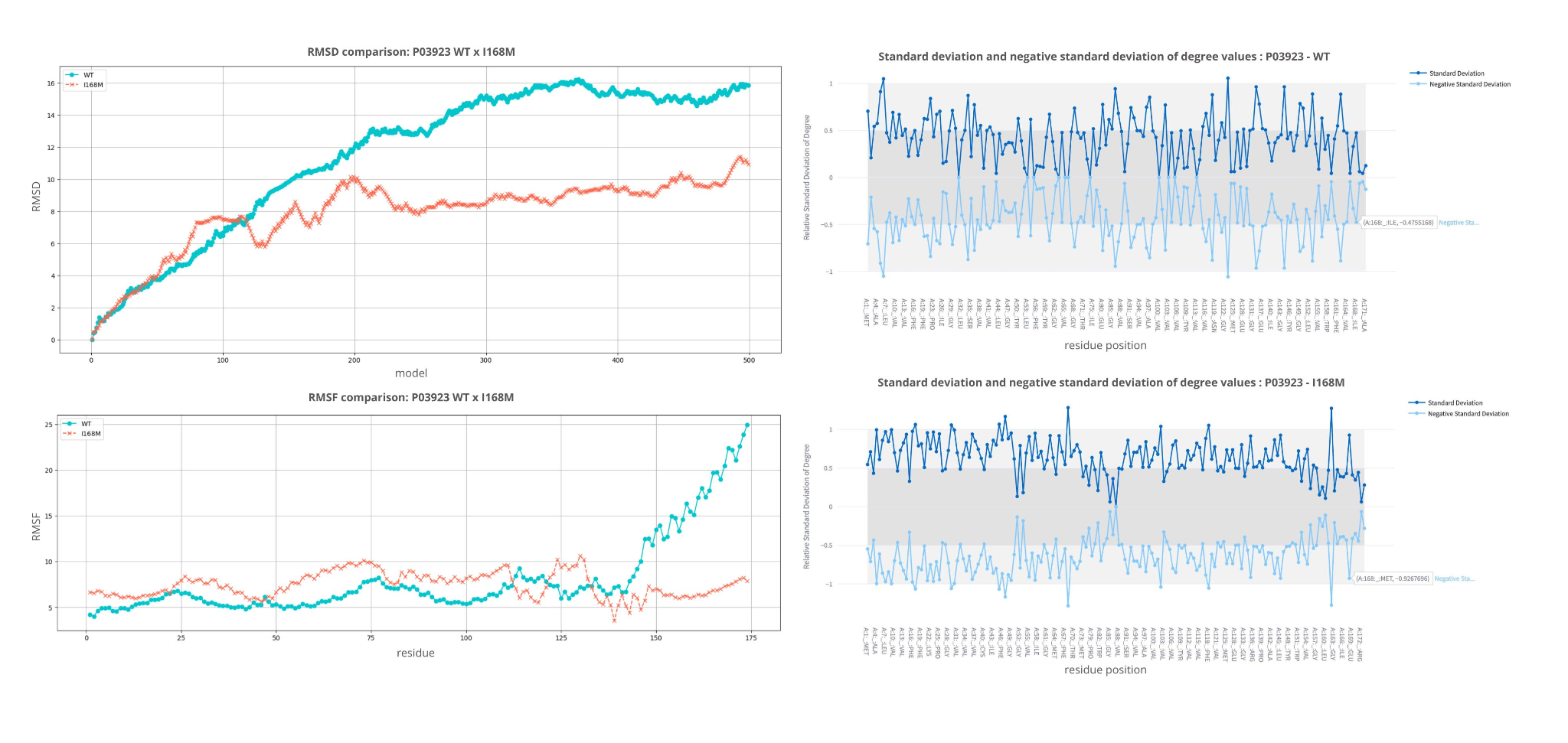
**Figure S12.** Structural comparison and analysis of dynamic variations of the P03923 protein, in the wild-type (WT) and mutant (I168M) forms. A: Comparison of RMSD values of the conformational variants along the simulated trajectory for WT (blue) and I168M (orange). B: Comparison of RMSF values per residue for WT (blue) and I168M (orange). C: Standard deviation and negative standard deviation of the number of bonds for P03923 (WT), evidencing variations in the stability of interactions observed in the conformational variants obtained in the dynamics. D: Standard deviation and negative standard deviation of the number of bonds for P03923 in the mutant form I168M, also demonstrating changes in the interaction patterns.
